# Plausibly Causal Plasma Proteins in Psychiatric Disorders: A Proteogenomic Appraisal

**DOI:** 10.64898/2026.07.10.26357744

**Authors:** Ruta Margelyte, Christina Dardani, Aimee L. Hanson, Xueyi Shen, Alexandra Havdahl, Dheeraj Rai, Andrew M. McIntosh, Naomi R. Wray, George Davey Smith, Gibran Hemani, Edward T. Bullmore, Tom R. Gaunt, Golam M. Khandaker

## Abstract

**Importance:** Large-scale plasma proteomic studies have linked hundreds of proteins to psychiatric disorders, but it is unclear which reflect causal biology rather than confounding or consequences of illness.

**Objective:** To determine which plasma proteins associated with depression, anxiety, bipolar disorder, and psychotic disorders have genetic evidence of a causal role.

**Design, Setting, and Participants:** This proteogenomic study combined genetic analyses with cross-sectional and longitudinal analyses in the UK Biobank (UKB) cohort (N=52,615). Associations of 2,923 plasma proteins were tested using two-sample Mendelian randomisation (MR) and genetic colocalisation with common-variant protein quantitative trait loci (pQTLs) from the UK Biobank Pharma Proteomics Project (N=34,557). Findings were appraised via a rule-based framework combining instrument validity with genetic analyses using blood and brain expression quantitative trait loci (eQTLs) and rare-variant MR.

**Exposures:** Measured and genetically proxied plasma protein levels.

**Main Outcomes and Measures:** Prevalent and incident diagnoses of depression, anxiety, bipolar disorder, and psychotic disorders; summary statistics from genome-wide association studies (GWAS) for depression, anxiety, bipolar disorder, and schizophrenia.

**Results:** A total of 1,685 proteins were involved in protein-disorder associations in UKB after multiple testing correction, of which 26 proteins (32 protein-disorder pairs) met both MR and colocalisation (MR false discovery rate <0.05 and posterior probability of colocalisation ≥0.8). After independent genetic validation, 9 disorder-protein pairs involving 7 proteins (CD40, ADAM22, CDHR1, PDIA3, SEMA3F, DSG3, and DDR1) met criteria for credible causal evidence, led by CD40, an immune signalling receptor, for schizophrenia and bipolar disorder. Specifically, higher genetically predicted CD40 levels were associated with lower risk of schizophrenia (odds ratio [OR] 0.90; 95%CI 0.85-0.95) and bipolar disorder (OR 0.89; 95%CI 0.84-0.95), supported by directionally concordant colocalised brain eQTL and rare-variant MR evidence.

**Conclusions and Relevance:** Few plasma protein associations with major psychiatric disorders had credible genetic support for causality, demonstrating that multi-layer appraisal across molecular layers, tissues, and genetic variant classes is required before nominating proteomic targets. CD40, an immune receptor targeted by drugs in clinical development, showed robust protective genetic support across schizophrenia and bipolar disorder, warranting focused mechanistic and therapeutic investigation.

**Key Points:** *Question:* Which plasma proteins associated with major psychiatric disorders have credible genetic evidence of a causal role?

*Findings:* In this proteogenomic study, 1,685 of 2,923 plasma proteins measured in 52,615 UK Biobank participants were associated with at least one psychiatric disorder. Of these, 26 proteins (32 protein-disorder pairs) were supported by Mendelian randomisation and colocalisation, with 7 proteins (9 pairs) meeting credible causal criteria after independent genetic validation.

*Meaning:* Multi-layer genetic appraisal narrowed 1,685 protein associations to a short priority list, led by CD40, an immune receptor targeted by drugs in clinical development, for schizophrenia and bipolar disorder.

## Introduction

Treatment options for major psychiatric disorders have changed little in decades, and many patients with depression or schizophrenia do not respond adequately to existing drugs[1, 2]. Drug targets with human genetic support are more likely to succeed in clinical development[3, 4], and circulating proteins are attractive candidates because they are measurable in blood and often pharmacologically tractable[5].

Large-scale plasma proteomics now measures thousands of proteins in population cohorts[6]. Associations with psychiatric disorders are, however, difficult to interpret as protein levels are strongly influenced by adiposity, smoking, socioeconomic factors, and illness itself, so an association may reflect confounding or a consequence of the disorder rather than a cause[7, 8]. Mendelian randomisation (MR), which uses genetic variants that influence protein levels as instruments, can address confounding and reverse causation, and genetic colocalisation tests whether a protein and a disorder share a causal gene[9, 10]. Proteome-wide MR studies have used these methods to nominate candidate proteins for psychiatric disorders variant[11, 12].

Two aspects of this evidence base limit what it can support. First, most findings rest on a single cis-acting variant, which precludes tests of heterogeneity and pleiotropy[13]. Second, many signals map to the major histocompatibility complex (MHC), where extensive linkage disequilibrium prevents attribution of an association to a specific gene[14], or rely on trans-acting variants that may act through other proteins. Most studies have relied on MR using plasma protein quantitative trait loci (pQTLs), with or without colocalisation[15, 16], and findings are rarely corroborated by independent genetic evidence from other molecular layers, tissues, or classes of variant[17].

We asked which plasma proteins associated with major psychiatric disorders are plausibly causal. Starting from candidate proteins prioritised through cross-sectional and longitudinal screening of 2,923 plasma proteins in UK Biobank, we conducted two-sample MR and colocalisation using common-variant pQTLs across four major psychiatric disorders: anxiety, depression, bipolar disorder, and schizophrenia. Every genetically supported finding was then evaluated using a rule-based appraisal framework that integrates instrument validity with independent validation across three genetic layers: MR with colocalisation using expression quantitative trait loci (eQTLs) in blood to tests whether genetic regulation of the encoding gene has a concordant effect and so guards against artifacts of protein measurement[18]; the same approach in brain cortex, a tissue directly relevant to psychiatric disorders[19]; and MR using rare variants, which are distinct from the common-variant instruments and, being in weak linkage disequilibrium with nearby variants, are less prone to confounding by neighbouring genes[20]. To our knowledge, this is the first study of major psychiatric disorders to systematically evaluate multi-layer genetic evidence across plasma proteins within a uniformly applied appraisal framework.

## Methods

### Study design

This study combined cross-sectional and longitudinal analyses of individual-level UK Biobank (UKB) data with genetic analyses using publicly available summary statistics (Figure 1). UKB has ethical approval from the NHS National Research Ethics Service North West (16/NW/0274). UKB data were accessed under application 26999. This work adhered to the STROBE and STROBE-MR reporting guidelines[21, 22]. A detailed description of the study methods is provided in eMethods in Supplement1.

**Figure 1.**
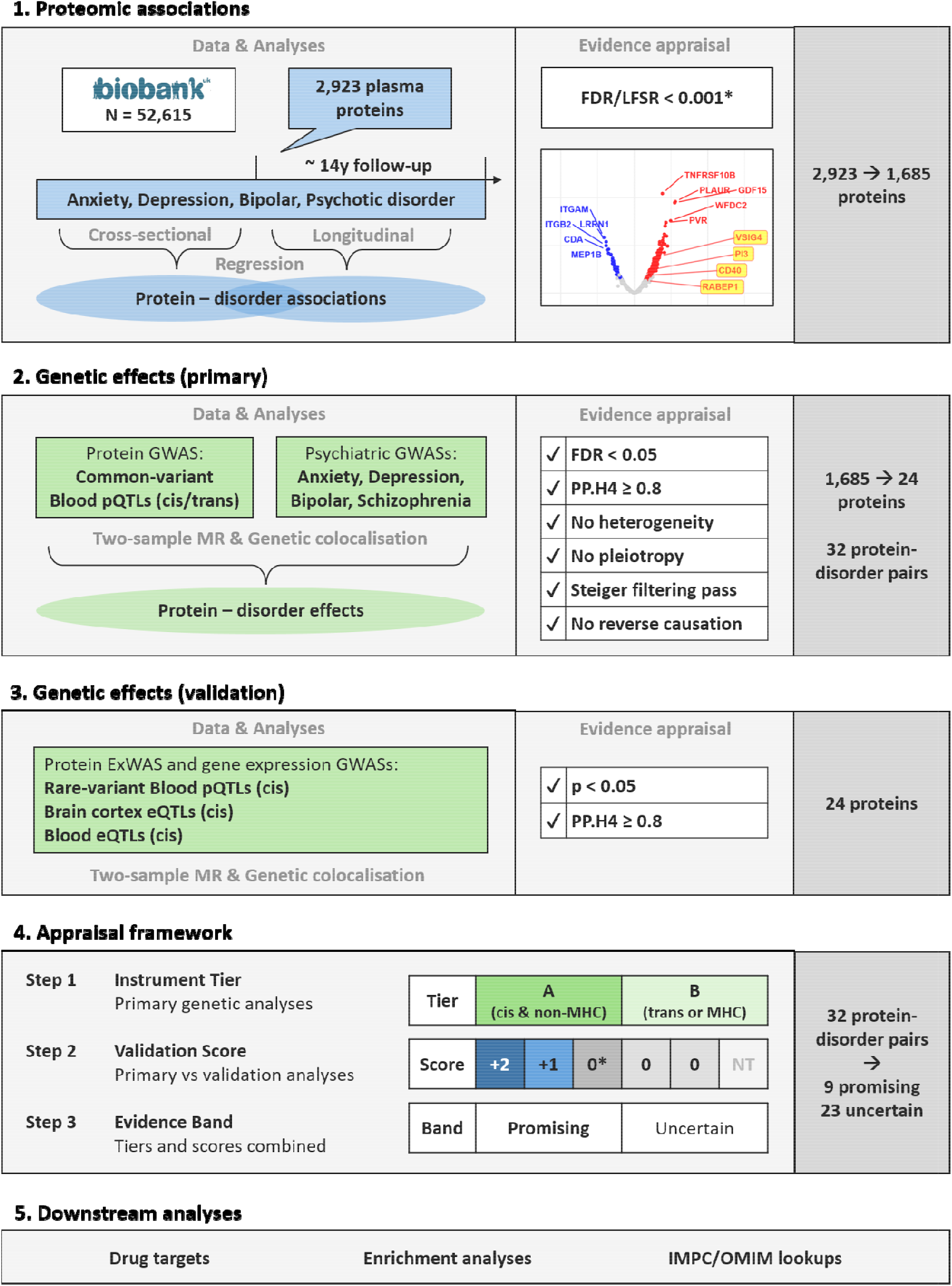
Study overview and analytical pipeline for identifying plausibly causal protein-disorder markers. (1) We examined associations between 2,920 plasma proteins and four psychiatric disorders in 52,615 UKB participants, both cross-sectionally at baseline (prevalent cases) and longitudinally at follow-up (incident cases), using logistic and Cox proportional hazards regression, respectively. A total of 1,685 proteins were involved in protein-disorder associations that were significant after multiple testing correction (FDR/LFSR < 0.001; * FDR/LFSR <0.05 for incident bipolar and psychotic disorders). (2) For protein-disorder associations identified in step (1), we performed two-sample MR and genetic colocalisation using common-variant blood pQTLs (cis-only, cis and trans, or trans-only) as genetic instruments for protein levels. A total of 32 protein-disorder markers across 26 proteins met all of the following criteria: MR FDR <0.05, genetic colocalisation PP.H4 ≥0.8, no evidence of heterogeneity or horizontal pleiotropy (where testable), passing Steiger filtering, and no evidence of reverse causation. (3) For proteins identified in step (2), we performed validation two-sample MR using rare or ultrarare variant blood pQTLs (cis-only), MR and genetic colocalisation using brain cortex eQTLs (cis-only), and MR and genetic colocalisation using blood eQTLs (cis-only). A validation analysis was considered significant if the MR P value was <0.05 and, for eQTL-based analyses, genetic colocalisation PP.H4 was ≥0.8. (4) Protein-disorder markers identified in step (2) were appraised in three steps. First, each marker was assigned a tier based on whether its common-variant instruments (pQTLs) could securely attribute the MR signal to the protein: Tier A (secure attribution; cis-acting instruments outside the MHC region) or Tier B (unresolved attribution; trans-acting instruments and/or instruments within the MHC region). Second, results from each validation analysis were compared with those of the primary analysis and scored as follows: +2 (significant and directionally concordant for the same disorder), +1 (significant and directionally concordant for a different disorder), 0* (significant and directionally discordant for the same disorder), 0 (all other results), or NT (not testable). Scores were summed across validation layers, with the number of layers tested also reported. Third, markers were classified into evidence bands based on their instrument tier and validation scores, with “Promising” markers having supporting evidence for a plausible causal protein-disorder association and all other markers classified as “Uncertain”. (5) We further characterised the proteins identified in steps (1) and (2) through functional enrichment analyses, assessment of therapeutic tractability (including information on clinical trials, drugs, and associated diseases), and a summary of reported human and mouse gene-phenotype associations. Abbreviations: eQTL, expression quantitative trait locus; ExWAS, exome-wide association study; FDR, false discovery rate; GWAS, genome-wide association study; IMPC, International Mouse Phenotyping Consortium; LFSR, local false sign rate; MHC, major histocompatibility complex; MR, Mendelian randomisation; OMIM, Online Mendelian Inheritance in Man; PP.H4, posterior probability of a shared causal variant; pQTL, protein quantitative trait locus; UKB, UK Biobank.

### Participants, proteomics, and outcomes in UK Biobank

UKB is a prospective population-based cohort of approximately 500,000 adults aged 40-69 years recruited in 2006-2010[23, 24]. Plasma levels of 2,923 proteins were measured using the Olink Explore 3072 platform in approximately 54,000 participants through the UKB Pharma Proteomics Project (UKB-PPP)[25, 26] (eTable1 in Supplement2). Participants missing >60% of proteins and proteins missing in >30% of participants were excluded. Remaining missing protein values were imputed using random forests[27]. Psychiatric outcomes included depression, anxiety, bipolar disorder, and psychotic disorders, identified through linked health records, death registry, and self-report. Prevalent cases had diagnoses recorded before or at baseline assessment, reflecting lifetime rather than current illness. Incident cases were first diagnosed during follow-up (eTable2 in Supplement2).

### Cross-sectional and longitudinal proteomic associations in UK Biobank

Associations of plasma protein levels with prevalent and incident disorders were estimated using logistic and Cox proportional hazards regression, respectively. Odds ratios (ORs) and hazard ratios (HRs), with 95% confidence intervals (CIs), were estimated per unit increase in normalised protein levels. Models were minimally adjusted for age, sex, and blood collection characteristics, and fully adjusted additionally for ethnicity, education, deprivation, body mass index, smoking, alcohol use, and physical activity (eTable2 in Supplement2).

Multiple testing was addressed using Benjamini-Hochberg false discovery rate (FDR) <0.001 and Empirical Bayes local false sign rate (LFSR) <0.001, the latter included to improve the true positive rate for rare outcomes[28–30]. For outcomes with frequency <1%, thresholds of 0.05 were also considered.

To assess specificity of the protein-disorder associations, selected non-psychiatric conditions served as external controls. Rheumatoid arthritis was the positive control, given its well-established inflammatory biology and expected immune-related proteomic signals[31]. Senile cataract and refraction and accommodation disorders were negative controls, as localised ocular conditions expected to show minimal or specific proteomic associations.

To identify over-represented biological pathways among proteins associated with psychiatric disorders, functional enrichment analyses were performed.

### Genetic causal inference using MR and colocalisation

Proteins associated with psychiatric disorders in the UKB were carried forward for genetic analyses. We performed two-sample bi-directional Mendelian randomisation (MR), using common-variant protein quantitative trait loci (pQTLs) as instrumental variables, to estimate the causal effect of genetically proxied protein levels on psychiatric outcomes[9, 32, 33]. Complementary pairwise conditional and colocalisation (PWCoCo) analyses evaluated shared causal variants at each locus, quantified by the posterior probability (PP.H4)[34–36].

Independent genome-wide significant cis- and trans-pQTLs (p ≤5×10^-8^; r² <0.001 within 10,000kb, F-statistics ≥10)[37] were selected from UKB European-ancestry GWAS (N=34,557)[25]. Outcome summary statistics were extracted from the largest available GWAS for anxiety (Cases=122,341, Controls=729,881), depression (Cases=294,322, Controls=741,438), bipolar disorder (Cases=41,917, Controls=371,549), and schizophrenia (Cases=76,755, Controls=243,649)[38–41].

Estimates were derived using the Wald ratio[42] or inverse-variance weighted (IVW) method[43]. Steiger filtering Cochran’s Q (heterogeneity), and MR-Egger intercept (directional pleiotropy) tests were performed where sufficient variants were available[44, 45]. Multiple testing was addressed using FDR correction[28]. In reverse MR, multiple testing was addressed using Bonferroni correction to account for the increased potential of pleiotropic influences when considering psychiatric exposures.

Protein-disorder pairs were considered potentially causal if they passed FDR (<0.05) and Steiger filtering, colocalised (PP.H4U≥U0.8), passed the heterogeneity and pleiotropy tests (where testable), and showed no evidence of reverse causation.

### Genetic validation

For each protein identified in the primary genetic analyses, we performed three validation analyses across four psychiatric disorders, each using cis-acting instruments: MR with colocalisation using expression quantitative trait loci (eQTLs) from blood (eQTLGen)[46] and brain cortex (MetaBrain)[19], and MR using rare (minor allele frequency [MAF] <1%) and ultrarare (MAF <0.1%) pQTLs identified in UKB exome-wide association study (ExWAS; N=49,736)[47],with psychiatric outcomes from a separate UKB ExWAS (N=454,787)[48]. A validation result was considered significant if the MR p-value was <0.05 and, for eQTL-based analyses, genetic colocalisation PP.H4 was ≥0.8.

### Appraisal framework

The appraisal framework (Table 1) assessed the causal plausibility of findings from the primary genetic analyses. MR and colocalisation identify a shared genetic signal but do not establish that the protein causes the disorder. The signal may act through a neighbouring gene, particularly in the major histocompatibility complex (MHC), or through trans-acting variants, and single-variant findings cannot be tested for pleiotropy. We therefore assessed how likely each instrument was to attribute the genetic effect on the disorder to the protein, and whether validation analyses corroborated the findings.

**Table 1:** Causal appraisal framework for protein-disorder markers. ^1^ Instrument tiers were assigned based on the characteristics of the instruments used in the primary analysis (1), which used MR and genetic colocalisation with common-variant blood pQTLs (cis-only or trans-only). ^2^ Validation scores were derived by comparing the result for each protein-disorder marker from the primary analysis (1) with the corresponding results from each validation analysis: (2) MR using rare or ultrarare variant blood pQTLs (cis-only), (3) MR and genetic colocalisation using brain cortex eQTLs (cis-only), and (4) MR and genetic colocalisation using blood eQTLs (cis-only). A validation analysis result was considered significant if it met following criteria: the MR P value was <0.05 and, for eQTL-based analyses, genetic colocalisation PP.H4 was ≥0.8. Directional concordance was assessed by comparing the signs of the primary and validation MR effect estimates (log odds ratios). “Same” indicates that the primary and validation analyses identified protein association with the same disorder; “Different” indicates that they identified protein associations with different disorder (among anxiety, depression, bipolar disorder, and schizophrenia). Abbreviations: eQTL, expression quantitative trait locus; MHC, major histocompatibility complex; MR, Mendelian randomisation; NT, not testable; PP.H4, posterior probability of a shared causal variant; pQTL, protein quantitative trait locus.

| Step 1: Instrument tier<sup>1</sup> |  |  |  |
|---|---|---|---|
| **Tier** | **Criterion** | **Interpretation** |  |
| A | Cis-acting instrument(s) only and<br>outside the MHC (chromosome 6, 25-34 Mb) | Attribution of the signal<br>to the protein is secure |  |
| B | Trans-acting instrument and/or<br>within the MHC (chromosome 6, 25-34 Mb) | Attribution of the signal<br>to the protein is unresolved |  |
| **Step 2: Validation score<sup>2</sup>** |  |  |  |
| **Score** | **Result<br>(validation)** | **Direction<br>(validation vs primary)** | **Disorder<br>(validation vs primary)** |
| +2 | Criteria met | Concordant | Same |
| +1 | Criteria met | Concordant | Different |
| 0* | Criteria met | Discordant | Same |
| 0 | Criteria met | Discordant | Different |
| 0 | Criteria not met | — | — |
| NT | No instrument available | — | — |
| **Step 3: Evidence band** |  |  |  |
| **Band** | **Rule** |  |  |
| **Promising** | Tier A instruments; One or more validation layers scoring +2;<br>Tier B instruments qualify only if the validation layers scoring +2 is from rare-variant MR. |  |  |
| Uncertain | All other findings |  |  |

In Step 1, instruments were classified as tier A (cis-acting, outside the extended MHC [chromosome 6, 25-34 Mb]), for which attribution to the protein is secure, or tier B (trans-acting or within the MHC), for which attribution is unresolved. In Step 2, validation evidence was scored by comparing each primary finding with the validation analyses. Scores reflected whether results met the evidence criteria, their direction of effect, and whether they concerned the same or a different disorder. Scores were summed across validation layers, with the number of layers tested also reported. In Step 3, findings based on tier A instruments were classified as promising if at least one validation result met the same-disorder evidence criteria. Findings based on tier B instruments were classified as promising only if rare-variant MR met those criteria. All other findings were classified as uncertain.

### Prior evidence and target characterisation

To distinguish replication from novel discoveries, our promising protein-disorder pairs were compared against prior MR and colocalisation studies of the same disorders via targeted literature searches. We further characterised all primary candidate proteins through an assessment of therapeutic tractability (including clinical trials, drugs, and disease indications)[49, 50] and a summary of reported human and mouse gene-phenotype associations[51, 52].

### Software

Analyses were conducted in R (v 4.4.2). Full details are available in eMethods in Supplement1.

## Results

A detailed description of the study results is provided in eResults in Supplement1.

### Cross-sectional and longitudinal proteomic associations in UK Biobank

Baseline data on 2,920 plasma protein analytes (eTable1 in Supplement2) were measured in 52,615 UK Biobank participants, median age of 58 years (range 40-70), 54% women, and 94% White ethnicity (eTable3 in Supplement2). At baseline, 6,085 participants (11.6%) had a lifetime diagnosis of at least one of the four psychiatric disorders, and 4,753 incident psychiatric diagnoses were recorded over a median follow-up of 13.2 years (interquartile range 12.4-14.0) (eTable4 in Supplement2).

After full covariate adjustment, we identified 830 protein-psychiatric disorder associations involving 574 proteins, with depression showing the broadest proteomic signature (424 proteins), followed by bipolar disorder (323), anxiety (45), and psychotic disorder (38). Adjusting beyond age, sex, and blood collection characteristics reduced associated proteins from 1,379 to 574, driven primarily by body mass index, smoking, and deprivation (eTable6 in Supplement2, eFigures2-5 in Supplement1). Across primary and sensitivity analyses, 1,685 proteins were associated with at least one psychiatric disorder (full results in eTable7 in Supplement2).

Plasma proteomics also captured disease-relevant signals in the negative and positive control conditions (eTable8 in Supplement2). Rheumatoid arthritis was associated with 779 proteins, dominated by immune markers. Senile cataract was associated with only 30 proteins, led by the lens protein β-crystallin B2 (CRYBB2), the sole protein associated with refraction and accommodation disorders.

### Genetic causal inference using MR and colocalisation

In primary genetic analyses, we identified 125 protein-psychiatric disorder pairs (90 plasma proteins) with MR estimates surviving multiple-testing correction at FDR <0.05. Of these, 32 pairs (26 proteins) showed genetic colocalisation (PP.H4 ≥0.8), passed Steiger filtering, and showed absence of heterogeneity and horizontal pleiotropy (where testable). 22 of these 26 proteins were instrumented by a single genetic variant. In reverse-direction analyses, genetic liability to depression showed causal effects on six plasma proteins after Bonferroni correction (N tests=6,764; p<7.4×10^-6^), none overlapping with the 26 primary proteins. Findings remained largely consistent in sensitivity analyses using depression and bipolar disorder GWAS excluding UKB participants. Genetic instruments are detailed in eTables11-12 (Supplement2), and primary MR and colocalisation results in eTables13-15 (Supplement2).

In validation genetic analyses of the 26 primary proteins across the four psychiatric disorders, five protein-disorder pairs showed supportive causal evidence in two-sample MR using rare or ultrarare genetic variants, while 10 pairs received additional support from brain and blood gene expression (eQTL) MR and colocalisation analyses.

Primary and validation genetic results are summarised in Figure 2 (and in an expanded version with cross-sectional and longitudinal UKB associations in eFigure11 in Supplement1). Validation instruments and results are detailed in eTables16-20 (Supplement2).

**Figure 2:**
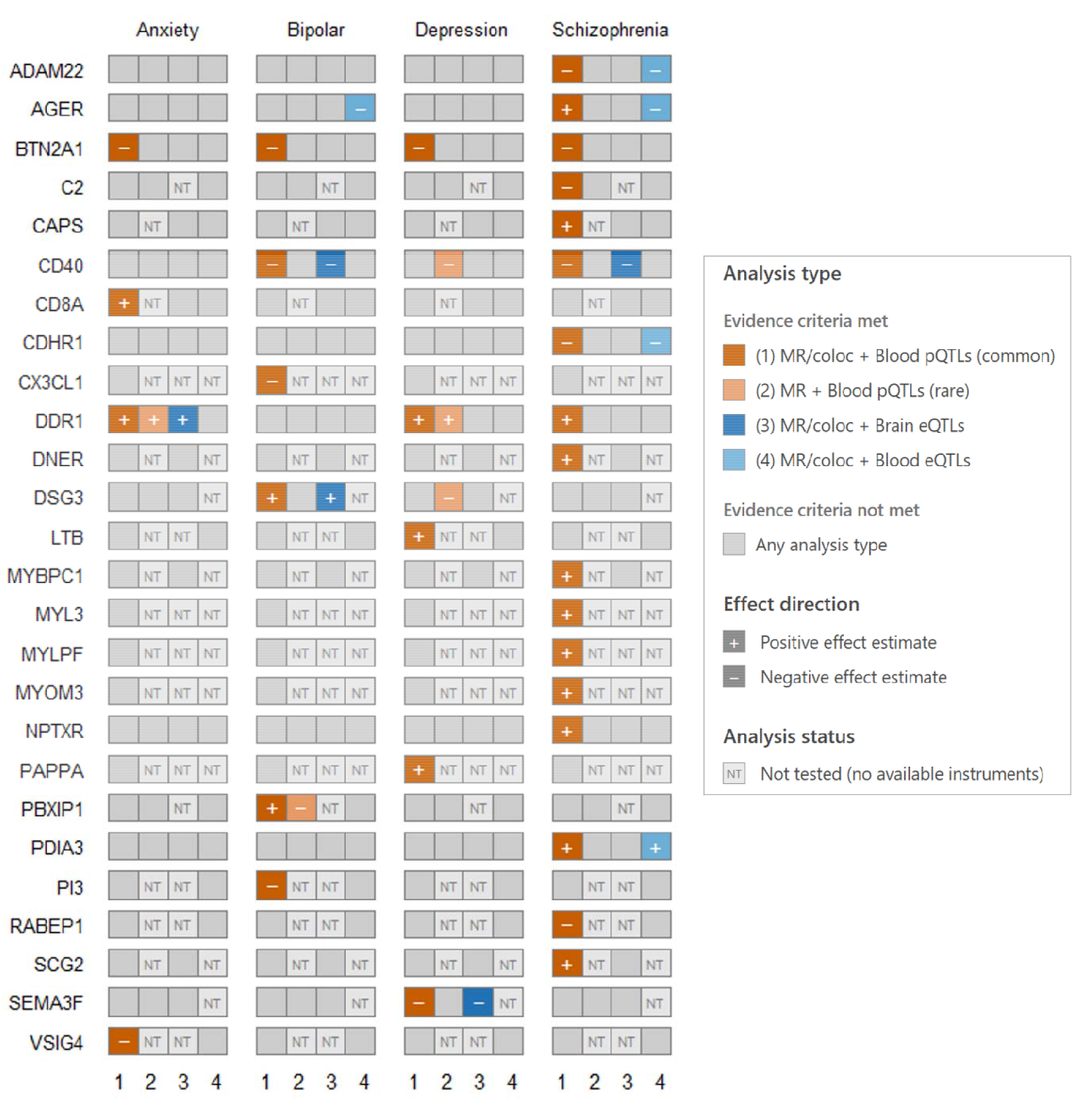
Genetic evidence for protein-disorder markers from primary and validation analyses. The primary analysis (1), using MR and genetic colocalisation with common-variant blood pQTLs (cis-only or trans-only), was considered to provide evidence for a protein-disorder association if it met all of the following criteria: MR FDR <0.05, genetic colocalisation PP.H4 ≥0.8, no evidence of heterogeneity or horizontal pleiotropy (where testable), passing Steiger filtering, and no evidence of reverse causation. 32 protein-disorder markers across 26 proteins met these criteria. The validation analyses comprised (2) MR using rare or ultrarare variant blood pQTLs (cis-only), (3) MR and genetic colocalisation using brain cortex eQTLs (cis-only), and (4) MR and genetic colocalisation using blood eQTLs (cis-only). A validation analysis was considered to provide supporting evidence if the MR P value was <0.05 and, for eQTL-based analyses, genetic colocalisation PP.H4 was ≥0.8. Effect direction was defined by the sign of the MR effect estimate (log odds ratio). Abbreviations: coloc, colocalisation; eQTL, expression quantitative trait locus, FDR, false discovery rate; MR, Mendelian randomisation; NT, not testable; PP.H4, posterior probability of a shared causal variant; pQTL, protein quantitative trait locus.

### Appraisal

All 32 primary protein-disorder pairs (26 proteins) were appraised by instrument tier, validation score, and final evidence band (Table 2, eTable21 in Supplement2). Of these, 9 pairs (7 proteins) had supporting evidence for a plausible causal association and were classified as “Promising,” with their primary MR estimates and variant counts detailed in Table 3.

**Table 2.**
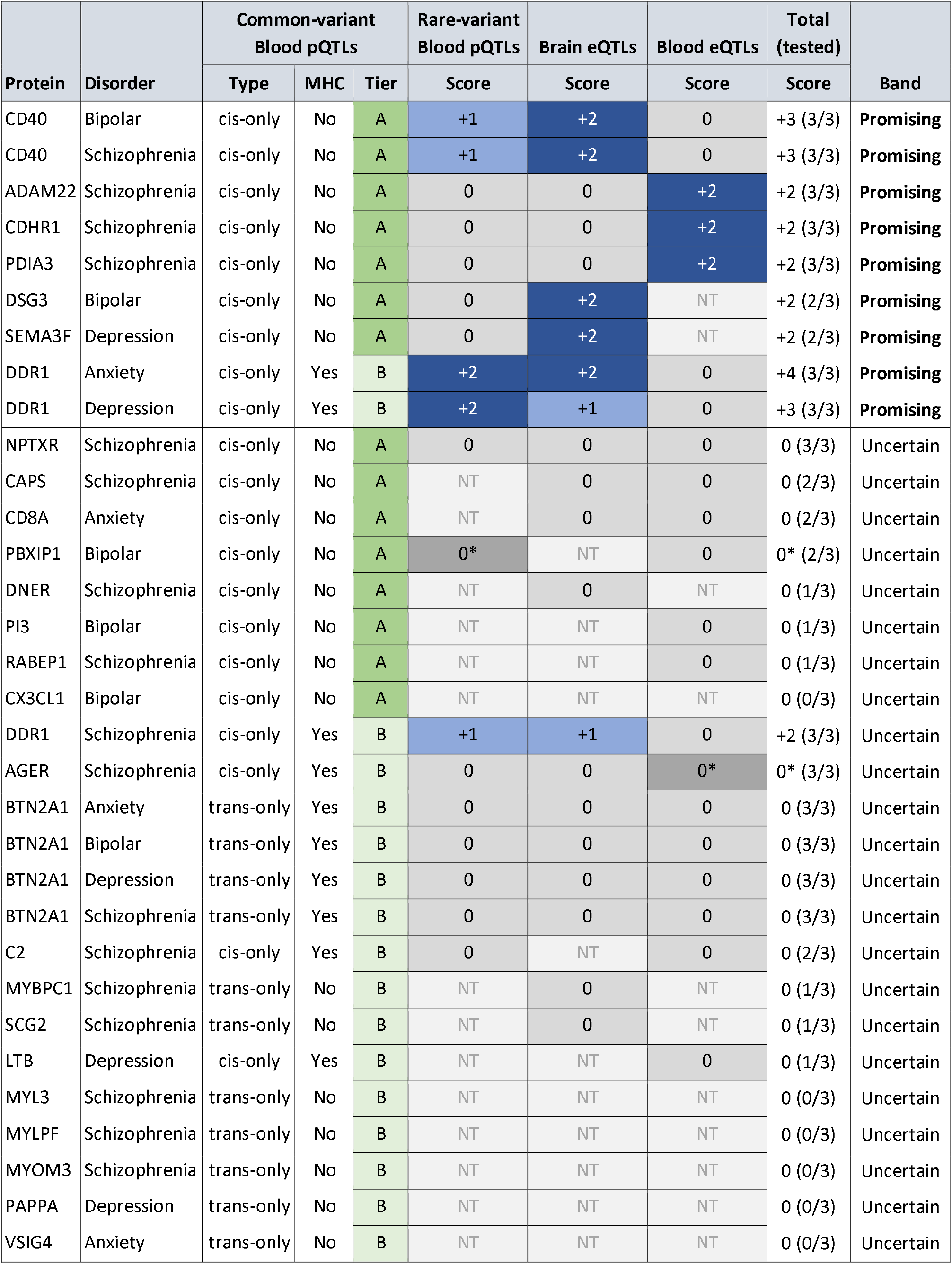
Causal appraisal of protein-disorder markers. 32 protein-disorder markers across 26 proteins from the primary analysis (1) using MR and genetic colocalisation with common-variant blood pQTLs (cis-only or trans-only) met the following criteria: MR FDR <0.05, genetic colocalisation PP.H4 ≥0.8, no evidence of heterogeneity or pleiotropy (where testable), passing Steiger filtering, and no evidence of reverse causation. These markers were eligible for causal appraisal. Each marker was assigned an instrument tier based on the ability of its pQTL instruments to attribute the MR signal to the protein: Tier A (secure attribution; cis-acting instruments outside the MHC region); Tier B (unresolved attribution; trans-acting instruments and/or instruments within the MHC region). Evidence for each marker was assessed independently across three validation layers: (2) MR using rare or ultrarare variant blood pQTLs (cis-only), (3) MR and genetic colocalisation using brain cortex eQTLs (cis-only), and (4) MR and genetic colocalisation using blood eQTLs (cis-only). A validation analysis was considered significant if the MR P value was <0.05 and, for eQTL-based analyses, genetic colocalisation PP.H4 was ≥0.8. Results from each validation analysis were compared with those of the primary analysis and scored as follows+2 (significant and directionally concordant for the same disorder), +1 (significant and directionally concordant for a different disorder), 0* (significant and directionally discordant for the same disorder), 0 (all other results), or NT (not testable). Scores were summed across validation layers, with the number of layers tested also reported. Markers were classified into evidence bands based on their instrument tier and validation scores, with “Promising” markers having supporting evidence for a plausible causal protein-disorder association and all other markers classified as “Uncertain”. Abbreviations: eQTL, expression quantitative trait locus; FDR, false discovery rate; MHC, major histocompatibility complex; MR, Mendelian randomisation; NT, not testable; PP.H4, posterior probability of a shared causal variant; pQTL, protein quantitative trait locus.

|  |  | Common-variant Blood pQTLs |  |  | Rare-variant Blood pQTLs | Brain eQTLs | Blood eQTLs | Total (tested) |  |
|---|---|---|---|---|---|---|---|---|---|
| Protein | Disorder | Type | MHC | Tier | Score | Score | Score | Score | Band |
| CD40 | Bipolar | cis-only | No | A | +1 | +2 | 0 | +3 (3/3) | **Promising** |
| CD40 | Schizophrenia | cis-only | No | A | +1 | +2 | 0 | +3 (3/3) | **Promising** |
| ADAM22 | Schizophrenia | cis-only | No | A | 0 | 0 | +2 | +2 (3/3) | **Promising** |
| CDHR1 | Schizophrenia | cis-only | No | A | 0 | 0 | +2 | +2 (3/3) | **Promising** |
| PDIA3 | Schizophrenia | cis-only | No | A | 0 | 0 | +2 | +2 (3/3) | **Promising** |
| DSG3 | Bipolar | cis-only | No | A | 0 | +2 | NT | +2 (2/3) | **Promising** |
| SEMA3F | Depression | cis-only | No | A | 0 | +2 | NT | +2 (2/3) | **Promising** |
| DDR1 | Anxiety | cis-only | Yes | B | +2 | +2 | 0 | +4 (3/3) | **Promising** |
| DDR1 | Depression | cis-only | Yes | B | +2 | +1 | 0 | +3 (3/3) | **Promising** |
| NPTXR | Schizophrenia | cis-only | No | A | 0 | 0 | 0 | 0 (3/3) | Uncertain |
| CAPS | Schizophrenia | cis-only | No | A | NT | 0 | 0 | 0 (2/3) | Uncertain |
| CD8A | Anxiety | cis-only | No | A | NT | 0 | 0 | 0 (2/3) | Uncertain |
| PBXIP1 | Bipolar | cis-only | No | A | 0* | NT | 0 | 0* (2/3) | Uncertain |
| DNER | Schizophrenia | cis-only | No | A | NT | 0 | NT | 0 (1/3) | Uncertain |
| PI3 | Bipolar | cis-only | No | A | NT | NT | 0 | 0 (1/3) | Uncertain |
| RABEP1 | Schizophrenia | cis-only | No | A | NT | NT | 0 | 0 (1/3) | Uncertain |
| CX3CL1 | Bipolar | cis-only | No | A | NT | NT | NT | 0 (0/3) | Uncertain |
| DDR1 | Schizophrenia | cis-only | Yes | B | +1 | +1 | 0 | +2 (3/3) | Uncertain |
| AGER | Schizophrenia | cis-only | Yes | B | 0 | 0 | 0* | 0* (3/3) | Uncertain |
| BTN2A1 | Anxiety | trans-only | Yes | B | 0 | 0 | 0 | 0 (3/3) | Uncertain |
| BTN2A1 | Bipolar | trans-only | Yes | B | 0 | 0 | 0 | 0 (3/3) | Uncertain |
| BTN2A1 | Depression | trans-only | Yes | B | 0 | 0 | 0 | 0 (3/3) | Uncertain |
| BTN2A1 | Schizophrenia | trans-only | Yes | B | 0 | 0 | 0 | 0 (3/3) | Uncertain |
| C2 | Schizophrenia | cis-only | Yes | B | 0 | NT | 0 | 0 (2/3) | Uncertain |
| MYBPC1 | Schizophrenia | trans-only | No | B | NT | 0 | NT | 0 (1/3) | Uncertain |
| SCG2 | Schizophrenia | trans-only | No | B | NT | 0 | NT | 0 (1/3) | Uncertain |
| LTB | Depression | cis-only | Yes | B | NT | NT | 0 | 0 (1/3) | Uncertain |
| MYL3 | Schizophrenia | trans-only | No | B | NT | NT | NT | 0 (0/3) | Uncertain |
| MYLPF | Schizophrenia | trans-only | No | B | NT | NT | NT | 0 (0/3) | Uncertain |
| MYOM3 | Schizophrenia | trans-only | No | B | NT | NT | NT | 0 (0/3) | Uncertain |
| PAPPA | Depression | trans-only | No | B | NT | NT | NT | 0 (0/3) | Uncertain |
| VSIG4 | Anxiety | trans-only | No | B | NT | NT | NT | 0 (0/3) | Uncertain |

**Table 3:** Genetic effects for promising protein-disorder markers from primary and validation analyses. Mendelian randomisation and genetic colocalisation results from primary and validation genetic analyses for prioritised protein-disorder markers. *** MR FDR<0.05 & colocalisation PP.H4>=0.8; ** MR p<0.05 & colocalisation PP.H4>=0.8; * MR p<0.05; Abbreviations: CI, confidence interval; eQTL, expression quantitative trait locus; FDR, false discovery rate; MR, Mendelian randomisation; PP.H4, posterior probability of a shared causal variant; OR, odds ratio; pQTL, protein quantitative trait locus.

| Protein | Disorder | Common variant<br>Blood pQTLs MR |  | Rare variant<br>Blood pQTLs MR |  | Brain eQTLs MR |  | Blood eQTLs MR |  |
|---|---|---|---|---|---|---|---|---|---|
|  |  | No. of variants | OR (95% CI) | No. of variants | OR (95% CI) | No. of variants | OR (95% CI) | No. of variants | OR (95% CI) |
| CD40 | Schizophrenia | 2 | **0.90 (0.85-0.95)\*\*\*** | 1 | 1.34 (0.22-8.23) | 1 | **0.89 (0.84-0.94)\*\*** | 2 | 0.95 (0.9-1.00)* |
| CD40 | Bipolar disorder | 2 | **0.89 (0.84-0.95)\*\*\*** | 1 | 0.60 (0.16-2.24) | 1 | **0.88 (0.82-0.93)\*\*** | 2 | 0.96 (0.88-1.05) |
| ADAM22 | Schizophrenia | 2 | **0.91 (0.87-0.96)\*\*\*** | 3 | 0.58 (0.13-2.53) | 1 | 0.99 (0.91-1.08) | 1 | **0.67 (0.45-0.89)\*\*** |
| CDHR1 | Schizophrenia | 1 | **0.86 (0.79-0.93)\*\*\*** | 1 | 0.14 (0.01-1.47) | 3 | 0.97 (0.94-1.00) | 2 | **0.93 (0.87-0.99)\*\*** |
| PDIA3 | Schizophrenia | 1 | **2.57 (1.69-3.91)\*\*\*** | 1 | 0.64 (0.09-4.47) | 1 | 1.20 (1.09-1.31)* | 1 | **1.45 (1.31-1.58)\*\*** |
| DSG3 | Bipolar disorder | 1 | **1.17 (1.08-1.26)\*\*\*** | 2 | 0.68 (0.25-1.89) | 1 | **1.25 (1.12-1.37)\*\*** | - | - |
| SEMA3F | Depression | 1 | **0.88 (0.84-0.93)\*\*\*** | 2 | 0.87 (0.44-1.73) | 1 | **0.96 (0.94-0.98)\*\*** | - | - |
| DDR1 | Anxiety | 1 | **1.07 (1.03-1.10)\*\*\*** | 3 | **1.19 (1.05-1.35)\*** | 1 | **1.15 (1.09-1.22)\*\*** | 4 | 0.99 (0.96-1.01) |
| DDR1 | Depression | 1 | **1.04 (1.02-1.06)\*\*\*** | 3 | **1.56 (1.05-2.33)\*** | 1 | 1.05 (1.00-1.11)* | 4 | 1.01 (0.98-1.04) |

CD40 was the top-scoring protein for schizophrenia and bipolar disorder, instrumented by two independent non-MHC cis variants (Tier A). Higher genetically predicted CD40 levels were associated with lower risk of schizophrenia (OR 0.90; 95%CI 0.85-0.95) and bipolar disorder (OR 0.89; 95%CI 0.84-0.95), supported by concordant brain cortex eQTL MR with colocalisation for schizophrenia (OR 0.89; 95%CI 0.84-0.94) and bipolar disorder (OR 0.88; 95%CI 0.83-0.93), and concordant rare-variant MR for depression (OR 0.54; 95%CI 0.40-0.74).

Five additional pairs with Tier A instruments were classified as “Promising” from a single validation layer scoring +2: ADAM22, CDHR1, and PDIA3 for schizophrenia (colocalised blood eQTL MR), and SEMA3F for depression and DSG3 for bipolar disorder (colocalised brain cortex eQTL MR). Two pairs instrumented by a single MHC cis-variant (Tier B), DDR1 for anxiety and depression, were also classified as “Promising” via concordant rare-variant MR (less vulnerable than common variants to the attribution ambiguity in the MHC), with DDR1-anxiety also supported by colocalised brain cortex eQTL MR. The remaining 23 pairs were classified as “Uncertain” due to non-significant or untestable validation layers, except for two directly contradicted pairs: AGER for schizophrenia (by colocalised blood eQTL MR) and PBXIP1 for bipolar disorder (by rare-variant MR).

### Discovery, replication, and target characterisation

Four of the nine promising pairs (CD40, ADAM22, CDHR1, and PDIA3, each for schizophrenia) replicated a prior proteome-wide MR and colocalisation study with closely matching estimates. CD40 for bipolar disorder has not been reported in a disorder-specific MR and colocalisation analysis but was implicated in cross-disorder studies[54, 55]. No prior MR and colocalisation reports were found for SEMA3F (depression), DSG3 (bipolar disorder), or DDR1 (anxiety or depression) (eTables27-28 in Supplement2).

Among the promising targets, human Mendelian phenotypes link ADAM22 to developmental and epileptic encephalopathy and CD40 to monogenic immune syndromes, supporting neurodevelopmental and immune processes in psychosis (eTable22 in Supplement2). Mouse knockout phenotypes link DDR1 to hyperactivity, supporting a role in behavioural regulation, and SEMA3F to behavioural and neurological abnormalities, including altered locomotor activity, impaired exploration, and abnormal sensory responses (eTable23 in Supplement2).

Of the 26 primary proteins, 17 are potentially or strongly druggable, and five have approved or clinical-stage drugs: CD40, LTB, and CX3CL1 (Phase II autoimmune trials), AGER (Phase III Alzheimer’s disease trial), and MYL3 (approved for cardiovascular indications) (eTables24-26 in Supplement2).

## Discussion

In this proteogenomic study, we combined cross-sectional and longitudinal population proteomics with causal inference approaches using common and rare genetic variation to systematically prioritise candidate causal proteins for major psychiatric disorders. More than half of the 2,923 proteins examined in UKB were associated with at least one psychiatric disorder, but only 26 proteins (32 protein-disorder pairs) met both MR and colocalisation criteria. Further genetic validation narrowed these to nine high-confidence pairs across seven proteins (CD40, ADAM22, CDHR1, PDIA3, SEMA3F, DSG3 and DDR1), led by the CD40 for schizophrenia and bipolar disorder.

The contrast between widespread proteomic associations and few genetically supported candidates reflects how strongly plasma protein levels are influenced by socioeconomic, behavioural, and health-related factors[8], complicating efforts to separate causal drivers from confounding or downstream effects[56]. Nevertheless, UKB control analyses for rheumatoid arthritis, senile cataract, and disorders of refraction and accommodation confirmed that plasma proteomics can still recover biologically meaningful disease signals despite these external influences. A key strength of our study is systematically validating genetic findings across molecular layers (protein abundance and gene expression), tissues (blood and brain), and variant classes (common and rare), overcoming the single-layer focus of previous proteome-wide psychiatric studies[15, 16, 53, 57, 58].

Among the prioritised proteins, CD40 is a tumour necrosis factor receptor superfamily member, expressed on antigen-presenting cells, regulates adaptive immune activation, and implicated in neuroinflammation[59]. CD40 showed consistent support across analyses, with colocalised effects of circulating blood protein levels on schizophrenia and bipolar disorder, based on two independent cis variants, and effects of brain cortex gene expression, alongside concordant rare-variant evidence for depression. These results align with prior MR studies and further implicate immune mechanisms in psychiatric disorders[60, 61]. Although agonist and antagonist CD40-targeting antibodies have entered clinical trials for cancer and autoimmune disease, the required direction of therapeutic modulation for psychiatric disorders remains unclear. Circulating CD40 largely comprises a soluble ectodomain shed from the cell surface, which can limit membrane-bound CD40 signalling[62]. Therefore, although higher genetically predicted circulating CD40 was associated with lower disorder risk, this could reflect greater receptor expression, increased shedding, or both, rather than increased CD40 signalling. Experimental work is needed to address these possibilities.

The remaining six prioritised proteins are plausible causal candidates, each with distinct biological support and caveats. ADAM22 (schizophrenia) and SEMA3F (depression) function in synaptic and axon-guidance pathways, with ADAM22 acting as the receptor for LGI1 in trans-synaptic signalling[63] and SEMA3F regulating limbic circuit development[64]. PDIA3 (schizophrenia) has independent transcriptomic, clinical, and experimental links to cognitive impairment[65]. DDR1 (anxiety and depression), a collagen-activated receptor tyrosine kinase implicated in oligodendrocyte signalling and blood-brain-barrier homeostasis[66–68]. However, its cis-acting variant instrument resides within the MHC region, where extensive linkage disequilibrium complicates causal attribution. For DSG3 (bipolar disorder), low bulk brain expression warrants cautious interpretation of its brain eQTL signal, although bulk data can mask cell-type-specific expression and peripheral mechanisms remain possible[69]. Finally, CDHR1 (schizophrenia), although primarily characterised by its role in retinal function, has also been associated with traumatic brain injury[70]. Overall, these candidates represent prioritised hypotheses for experimental follow-up.

## Limitations

Although applied uniformly using explicit rules, our appraisal framework relies on analytical judgements, and alternative thresholds could reclassify borderline findings. Second, most genetic instruments were single variants, precluding heterogeneity and pleiotropy sensitivity analyses, a limitation we sought to address through genetic validation analyses. Third, plasma protein levels may not reflect brain biology, and eQTL data were limited to bulk cortex, with several proteins lacking brain instruments. However, bidirectional peripheral-central immune and neurovascular communication[71, 72] and shared blood-brain molecular regulation[73] support the relevance of blood-based findings to brain phenotypes. Fourth, the anxiety, depression, and bipolar disorder GWAS included UKB participants, and rare-variant analyses were conducted within the UKB, meaning sample overlap could bias estimates toward phenotypic associations. Fifth, in UKB analyses, diagnoses relied on healthcare records and self-report, risking misclassification. Bipolar disorder and schizophrenia cases were few, limiting statistical power, while the predominantly middle-aged, European-ancestry cohort limits generalisability. Finally, two candidate proteins meeting primary genetic criteria, AGER (schizophrenia) and PBXIP1 (bipolar disorder), showed directionally opposite effects in validation analyses, and were not prioritised, although such discordance could reflect distinct regulatory mechanisms rather than a lack of causal biology.

## Conclusions

Few plasma protein associations with major psychiatric disorders showed credible genetic support for a causal role. Of seven prioritised proteins, CD40 had the strongest evidence, specifically for schizophrenia and bipolar disorder, warranting follow-up to clarify therapeutic directionality. While plasma proteomics can nominate causal candidates, triangulation across molecular layers, tissues, and variant classes is essential to establish causal support.

## Article Information

### Author Contributions

R.M., C.D., and G.M.K were responsible for the conception and design of the study. R.M. was responsible for UKB data preparation and cleaning. C.D. conducted preparation of genetic data. R.M. and C.D. carried out data analyses and were responsible for the drafting of the original manuscript. All co-authors critically reviewed and revised the manuscript and approved the final version.

## Data Sharing Statement

UK Biobank data are available to bona fide researchers on application (https://www.ukbiobank.ac.uk). Data supporting the results of the present study are available from the UKB (https://www.ukbiobank.ac.uk/enable-your-research/apply-for-access) to researchers with UKB approval.

Genome-wide summary statistics are publicly available from the sources cited. UKB blood pQTL data: http://ukb-ppp.gwas.eu

Blood eQTL data: https://www.eqtlgen.org/phase1.html

Brain cortex eQTL data at MetaBrain platform: https://www.metabrain.nl/

GWAS data on anxiety, bipolar, and schizophrenia: https://pgc.unc.edu/for-researchers/download-results/

GWAS data on depression: https://ipsych.dk/en/research/downloads/

ExWAS data (psychiatric disorders): https://www.ebi.ac.uk/gwas/publications/34662886

Therapeutic tractability and clinical trials data at the Open Targets Platform: https://platform.opentargets.org/

GO annotations: https://geneontology.org/docs/go-annotations/

Tissue expression at The Human Protein Atlas: https://www.proteinatlas.org/

KEGG pathways: https://www.genome.jp/kegg/

REACTOME pathways: https://reactome.org/

IMPC annotations: https://www.mousephenotype.org/

OMIM annotations: https://www.omim.org/

Analytical code is available at https://github.com/rutamargelyte/proteomics_psychiatric_analyses.

## Additional Contributions

We thank the participants and staff of UK Biobank. Data were accessed under UK Biobank application 26999.

## Data Availability

All data produced in this study are provided in the supplementary materials accompanying the manuscript. The underlying source data are available through public repositories or established controlled-access procedures.

http://ukb-ppp.gwas.eu

https://www.synapse.org/Synapse:syn23627957

https://www.eqtlgen.org/phase1.html

https://www.metabrain.nl/

https://pgc.unc.edu/for-researchers/download-results/

https://ipsych.dk/en/research/downloads/

https://www.ebi.ac.uk/gwas/publications/34662886

https://platform.opentargets.org/

https://www.mousephenotype.org/

https://www.omim.org/

