## Supplement1 for "Plausibly Causal Plasma Proteins in Psychiatric Disorders: A Proteogenomic Appraisal"

eMethods

Participants, proteomics, and outcomes in UK Biobank

UKB is a prospective population-based cohort of approximately 500,000 adults aged 40-69 years recruited in 2006-2010[1, 2]. At baseline, participants provided written informed consent, completed detailed questionnaires on sociodemographic factors, lifestyle, and medical history, underwent physical and anthropometric assessments, and provided biological samples. UKB combines rich phenotypic data with genome-wide genotyping, exome sequencing, biomarkers assays, imaging data, and linkage to routinely collected electronic health records, with ongoing follow-up for health outcomes.

Plasma levels of 2,923 proteins were measured using the Olink Explore 3072 platform in approximately 54,000 participants through the UKB Pharma Proteomics Project (UKB-PPP)[3, 4] (eTable1 in Supplement2). The UKB-PPP includes ~54,000 participants, of whom 86% were randomly selected from baseline, 12% were preselected by consortium members based on certain characteristics of interest, and 2% included as a part of a COVID-19 study. Participants with baseline proteomic data from the randomised or consortium-selected subsets were included in the present analyses. A total of 2,923 plasma proteins were measured across eight panels: Cardiovascular, Cardiovascular II, Inflammation, Inflammation II, Neurology, Neurology II, Oncology, and Oncology II. Protein abundance was expressed as Normalized Protein eXpression (NPX) values on the log₂ scale, corrected for plate and batch. Details of the Olink proteomics assay, data processing and quality control are reported in previous publications[4]. Participants missing >60% of proteins and proteins missing in >30% of participants were excluded. Remaining missing protein values were imputed using random forests[5]. Imputation was performed separately for each panel and including participants’ age and sex as predictors, with 50 trees per forest grown using subsampling without replacement.

Psychiatric outcomes included depression, anxiety, bipolar disorder, and psychotic disorders, identified through linked health records, death registry, and self-report. First-occurrence fields in UKB were used to determine the earliest recorded date of diagnosis across data sources. Prevalent cases had diagnoses recorded before or at baseline assessment, reflecting lifetime rather than current illness. Incident cases were first diagnosed during follow-up (eTable2 in Supplement2). Outcomes were analysed as binary variables. Follow-up for incident outcomes began at the date of baseline assessment and continued until the earliest of (1) first diagnosis of the outcome of interest, (2) death, or (3) the last available date of hospital inpatient records. Death was treated as a censoring event. Time to first diagnosis or censoring was calculated in months.

Cross-sectional and longitudinal proteomic associations in UK Biobank

Associations of plasma protein levels with prevalent and incident disorders were estimated using logistic and Cox proportional hazards regression, respectively. Odds ratios (ORs) and hazard ratios (HRs), with 95% confidence intervals (CIs), were estimated per unit increase in normalised protein levels. To distinguish proteins potentially associated with disorder onset or disorder state, proteins were classified as associated with incident-only, prevalent-only, or both incident and prevalent disorder. Participants with CRP >10 mg/L were excluded to reduce the effect of concurrent infection on the plasma proteome[6]. In cross-sectional analyses, individuals with other psychiatric diagnoses were excluded from controls to improve case-control contrast, by avoiding contamination with closely related phenotypes, thereby estimating protein associations distinguishing psychiatric cases from psychiatrically well participants. In longitudinal analyses, participants with the outcome at baseline and those censored within 12 months were excluded to minimise reverse causation.

Models were minimally adjusted for age, sex, and blood collection characteristics, and fully adjusted additionally for ethnicity, education, deprivation, body mass index, smoking, alcohol use, and physical activity (eTable2 in Supplement2). Potential confounders were selected a priori based on data availability in UK Biobank. Socio-demographic variables included age, sex, Townsend deprivation index (TDI), highest educational attainment (higher, secondary, or vocational/other), and self-reported ethnicity (White or Other). Lifestyle-related factors included body mass index (BMI), smoking status (never, previous, or current), alcohol consumption (current or previous/never), and physical activity level (low, moderate, or vigorous). Variables related to blood protein measurement included fasting duration at blood collection, season of sample collection (spring, summer, autumn, or winter), and hour of assessment. All covariates were obtained at baseline assessment. . Missing covariate data were handled using multiple imputation by chained equations (MICE), with estimates pooled across imputed datasets using Rubin’s rules[7, 8].

Multiple testing was addressed using Benjamini-Hochberg false discovery rate (FDR) <0.001 and Empirical Bayes local false sign rate (LFSR) <0.001, the latter included to improve the true positive rate for rare outcomes[9-11]. For outcomes with frequency <1%, thresholds of 0.05 were also considered.

To assess specificity of the protein-disorder associations, selected non-psychiatric conditions served as external controls. Rheumatoid arthritis was the positive control, given its well-established inflammatory biology and expected immune-related proteomic signals[12]. Senile cataract and refraction and accommodation disorders were negative controls, as localised ocular conditions expected to show minimal or specific proteomic associations.

To assess the robustness of the primary findings, models were repeated using unimputed protein data, including participants with CRP >10 mg/L, and applying stricter definitions of depression and anxiety based solely on health records (excluding self-report).

To identify over-represented biological pathways among proteins associated with psychiatric disorders, functional enrichment analyses were performed. Proteins from fully covariate adjusted models were taken forward for Reactome and Kyoto Encyclopaedia of Genes and Genomes (KEGG) pathway enrichment, stratified (where appropriate) into proteins associated in incident only, prevalent only, and both incident and prevalent disorders[13, 14]. Enrichment was also assessed for proteins associated with three or more psychiatric disorders. Genes encoding the 2,920 UKB proteins were used as the background. Significance was defined using FDR <0.05, with nominal p <0.05 also considered.

Genetic causal inference using MR and colocalisation

Proteins associated with psychiatric disorders in the UKB cross-sectional and longitudinal analyses were carried forward for genetic analyses.

We performed two-sample bi-directional Mendelian randomisation (MR), using common-variant protein quantitative trait loci (pQTLs) as instrumental variables, to estimate the causal effect of genetically proxied protein levels on psychiatric outcomes[15-17]. Complementary pairwise conditional and colocalisation (PWCoCo) analyses evaluated shared causal variants at each locus, quantified by the posterior probability (PP.H4)[18-20].

Independent genome-wide significant cis- and trans- pQTLs (p ≤5×10^-8^; r² <0.001 within 10,000kb, F-statistics ≥10)[37] were selected from UKB European-ancestry GWAS (N=34,557)[4]. Both cis and trans genetic instruments were available: cis variants were defined was those located within a 1 Mb window around the gene, and trans variants was those located outside this window. Cis variants are more likely to directly influence gene expression and protein levels and are therefore less prone to horizontal pleiotropy[21]. The inclusion of trans variants may increase the proportion of variance explained in protein levels and improve statistical power but increase the risk of pleiotropy or genetic confounding[22, 23]. Therefore, analyses were conducted separately using specific (cis-only) and broader (cis + trans or trans only) instrument sets. Outcome summary statistics of single nucleotide polymorphisms (SNPs) associated with psychiatric disorders were extracted from the largest available GWAS for anxiety (Cases=122,341, Controls=729,881), depression (Cases=294,322, Controls=741,438), bipolar disorder (Cases=41,917, Controls=371,549), and schizophrenia (Cases=76,755, Controls=243,649)[24-27]. The SNP-exposure and SNP-outcome alleles were harmonised to ensure that effects correspond to the same reference allele. To assess potential bias due to sample overlap with the UKB proteomic GWAS, we additionally used depression and bipolar disorder GWAS summary statistics excluding UK Biobank participants (depression: 166,773 cases and 507,679 controls; bipolar disorder: 40,463 cases and 313,436 controls).

Estimates were derived using the Wald ratio[28] or inverse-variance weighted (IVW) method[29]. Steiger filtering Cochran’s Q (heterogeneity), and MR-Egger intercept (directional pleiotropy) tests were performed where sufficient variants were available[30, 31]. We applied Steiger filtering to prioritise genetic instruments explaining more variance in the exposure than the outcome, where variants explaining greater variance in the outcome were excluded, as this suggests that the primary phenotype influenced by the variant may be the psychiatric outcome rather than the protein abundance. Multiple testing was addressed using FDR correction[9].

Colocalisation analyses were performed to assess whether the genetic association signals for protein levels and psychiatric outcomes at a given locus were driven by the same variant or by distinct variants in linkage disequilibrium (LD), thereby complementing MR analyses[18, 19]. For colocalisation analyses, we prioritised variants with the greatest biological relevance to the protein. Specifically, (1) for instruments containing at least one cis variant, cis variants were selected; (2) for instruments comprising multiple trans variants only, the variant with the smallest p-value was selected to minimise potential pleiotropic effects; and (3) single-variant instruments were tested regardless of whether they were cis or trans. We extracted regions within ±500 kb around each instrumented variant and performed PWCoCo analysis to compare all conditionally independent signals in the exposure and outcome datasets[20]. We used genotype data from mothers in the Avon Longitudinal Study of Parents and Children (ALSPAC; N = 7,733) as the LD reference panel[32]. Evidence of colocalisation was defined as a posterior probability for H4 (PPH4) ≥ 0.80, indicating strong support for a shared causal variant. Variants located within the Major Histocompatibility Complex (MHC) were interpreted cautiously due to the complex LD structure of this region hindering resolution of distinct causal signals[33].

To assess reverse causation, we performed bi-directional MR analyses to evaluate if genetic liability to psychiatric disorders influenced circulating protein levels. Genetic instruments for psychiatric disorders were selected at genome-wide significance (p ≤ 5 × 10⁻⁸) and were independent (r² < 0.001; 10,000 kb window). Selected instruments for psychiatric outcomes and proteins were harmonised, and causal effects were estimated using the IVW method. In reverse MR, multiple testing was addressed using Bonferroni correction to account for the increased potential of pleiotropic influences when considering psychiatric exposures

Protein-disorder pairs were considered potentially causal if they passed FDR (<0.05) and Steiger filtering, colocalised (PP.H4 ≥ 0.8), passed the heterogeneity and pleiotropy tests (where testable), and showed no evidence of reverse causation.

Genetic validation

For each protein identified in the primary genetic analyses, we performed three validation analyses across four psychiatric disorders, each using cis-acting instruments: MR with colocalisation using expression quantitative trait loci (eQTLs) from blood (eQTLGen)[34] and brain cortex (MetaBrain)[35], and MR using rare (minor allele frequency [MAF] <1%) and ultrarare (MAF <0.1%) pQTLs identified in UKB exome-wide association study (ExWAS; N=49,736)[36],with psychiatric outcomes from a separate UKB ExWAS (N=454,787)[37].

In the two-sample MR analyses, we used genetic instruments that were genome-wide significant (5 × 10^-8^) and approximately independent (r2<0.1). For each exposure and each psychiatric outcome, we harmonised variant-exposure and variant-outcome alleles to ensure that effect estimates corresponded to the same reference allele. When a protein was instrumented by a single variant, causal estimates were derived using the Wald ratio method[28]. When two or more independent variants were available, inverse-variance weighted (IVW) regression was applied as the primary analysis[29].

MR analyses using rare and ultrarare variants were complemented with weighted median, weighted mode, and simple mode methods to provide robustness against invalid instruments and identify consistent causal signals when some variants violate IV assumptions. Rare protein-coding variants typically have larger effect sizes and clearer functional consequences compared to common variants, that may lie in non-coding regions and could be confounded by linkage disequilibrium (LD). Consistent effects across analyses using common and rare variants increase confidence that the effect is driven by the protein itself[38, 39].

A validation result was considered significant if the MR p-value was <0.05 and, for eQTL-based analyses, genetic colocalisation PP.H4 was ≥0.8.

Appraisal framework

The appraisal framework (Table 1) assessed the causal plausibility of findings from the primary genetic analyses. MR and colocalisation identify a shared genetic signal but do not establish that the protein causes the disorder. The signal may act through a neighbouring gene, particularly in the major histocompatibility complex (MHC), or through trans-acting variants, and single-variant findings cannot be tested for pleiotropy. We therefore assessed how likely each instrument was to attribute the genetic effect on the disorder to the protein, and whether validation analyses corroborated the findings.

In Step 1, instruments were classified as tier A (cis-acting, outside the extended MHC [chromosome 6, 25-34 Mb]), for which attribution to the protein is secure, or tier B (trans-acting or within the MHC), for which attribution is unresolved. In Step 2, validation evidence was scored by comparing each primary finding with the validation analyses. Scores reflected whether results met the evidence criteria, their direction of effect, and whether they concerned the same or a different disorder. Scores were summed across validation layers, with the number of layers tested also reported. In Step 3, findings based on tier A instruments were classified as promising if at least one validation result met the same-disorder evidence criteria. Findings based on tier B instruments were classified as promising only if rare-variant MR met those criteria. All other findings were classified as uncertain.

Prior evidence and target characterisation

To distinguish replication from novel discoveries, our promising protein-disorder pairs were compared against prior MR and colocalisation studies of the same disorders via targeted literature searches. We further characterised all primary candidate proteins through an assessment of therapeutic tractability (including clinical trials, drugs, and disease indications)[40, 41] and a summary of reported human and mouse gene-phenotype associations[42, 43].

Data on the potential therapeutic tractability of protein targets were retrieved for any treatment modality (small molecule, antibody binding, and/or others) from the Open Targets Platform. The Open Targets Platform (<https://platform.opentargets.org/>) is a publicly available resource for drug target identification and prioritisation that integrates genetic and genomic evidence with data on protein structure, function, approved drugs, and ongoing clinical trials. Open Targets categorises target tractability using predefined groups (buckets): eight for small molecules and nine for antibodies (<https://github.com/chembl/tractability_pipeline_v2>). To aid interpretation, we grouped these buckets into three mutually exclusive categories, consistent with previous work[44]: (1) strong druggability evidence (buckets 1-3 for small molecules, antibodies, and other modalities), (2) likely or potentially druggable (buckets 4-8 for small molecules and 4-5 for antibodies), and (3) little or unknown druggability evidence (remaining buckets). Data were retrieved on 26/05/2026.

We used the Online Mendelian Inheritance in Man (OMIM) database to identify known links between the encoding genes for identified proteins and human disease. OMIM provides detailed, referenced summaries of known Mendelian conditions and associated genes, with a current catalogue exceeding 16,000 genes. Furthermore, we used data from the International Mouse Phenotyping Consortium (IMPC) to assess phenotypic consequences of gene disruption in experimental models. The IMPC systematically generates and characterises knockout mouse models, providing standardised high-throughput phenotyping data that link gene disruption to observable traits and potential disease relevance. Data were retrieved on 26/05/2026.

Software

Analyses were conducted in R (v 4.4.2).

Cross-sectional and longitudinal cohort analyses used R packages stats (v4.4.2), survival (v4.4.2), lspline (1.0-0), and ashr (v2.2-63).

Missing data imputation was done with R packages mice (v3.18.0)[45] and missForest (v1.5)[5].

Enrichment analyses were performed with R packages ReactomePA (v1.50.0) and clusterProfiler (v4.14.6)[46].

Genetic analyses: Blood plasma pQTL data and blood cell derived eQTL data were extracted and processed using R package gwasvcf (v1.0) (<https://github.com/MRCIEU/gwasvcf>)[47]. The summary data from MetaBrain were lifted over from GRCh38 to GRCh37 using the UCSC liftover tool to match the build of the rest of the data[48]. Two-sample MR, Steiger filtering, and bi-directional MR analyses were conducted using functions from R packages TwoSampleMR (v0.5.6) (<https://github.com/MRCIEU/TwoSampleMR>) and mrpipeline (v1.0) (<https://github.com/jwr-git/mrpipeline>)[49]. The PWCoCo algorithm was implemented using the Pair-Wise Conditional analysis and Colocalisation analysis package v1.0 (<https://github.com/jwr-git/pwcoco>)[20].

Data on the therapeutic tractability, known drugs, and clinical trials were extracted using functions from R package otargen (v2.0.1)[50].

Gene Ontology (GO) and OMIM annotations were retrieved using R package biomaRt (v2.62.1).

Mouse knockout phenotype data were obtained from the IMPC database via its public API. Human-to-mouse orthologs were mapped using R package homologene (v1.48.6).

Data was visualised using R package ggplot2 (v4.0.1).

Genetic analyses and protein imputation were carried out using the computational facilities of the Advanced Computing Research Centre of the University of Bristol (<http://www.bris.ac.uk/acrc/>).

eResults

Cross-sectional and longitudinal proteomic associations in UK Biobank

Baseline data on 2,920 plasma protein analytes (eTable1 in Supplement2) were measured in 52,615 UK Biobank participants, median age of 58 years (range 40-70), 54% women, and 94% White ethnicity (eTable3 in Supplement2). Participant and protein selection details are presented in eFigure1 in Supplement1.

At baseline, 6,085 participants (11.6%) had a lifetime diagnosis of at least one of the four psychiatric disorders, comprising 7,211 diagnoses across depression (n=4,655; 8.9%), anxiety (n=2,158; 4.1%), bipolar disorder (n=217; 0.4%), and psychotic disorder (n=181; 0.3%), and 4,753 incident psychiatric diagnoses were recorded over a median follow-up of 13.2 years (interquartile range 12.4-14.0), comprising largely depression (n=2,418; 5.0%) and anxiety (n=2,863; 5.7%), and relatively fewer cases of bipolar disorder (n=105; 0.2%) and psychotic disorder (n=172; 0.3%) (eTable4 in Supplement2). Psychiatric comorbidity (lifetime co-diagnosis) was common, with up to 18.5% of participants having more than one psychiatric diagnosis at baseline (eTable5 in Supplement2).

To identify psychiatric disorder relevant plasma proteins, we examined the cross-sectional and longitudinal associations of circulating levels of 2,920 proteins with prevalent and incident psychiatric disorders, respectively, using logistic regression and Cox proportional hazards models. Primary analyses included 47,730 participants after excluding those with CRP >10 mg/ L (n=2,180; 4.2%), indicative of acute infection, and those with missing CRP data (n=2,705; 5.1%). To account for multiple testing, we applied significance thresholds of local false sign rate (LFSR) or false discovery rate (FDR) <0.001 to analysis of each prevalent disorder and to analysis of each incident disorder except for bipolar and psychotic disorders, which were tested with a less stringent threshold (LFSR/FDR <0.05) due to the smaller number of incident diagnoses.

After full covariate adjustment, we identified 830 protein-psychiatric disorder associations involving 574 proteins, with depression showing the broadest proteomic signature (424 proteins), followed by bipolar disorder (323), anxiety (45), and psychotic disorder (38). Adjusting beyond age, sex, and blood collection characteristics reduced associated proteins from 1,379 to 574, driven primarily by body mass index, smoking, and deprivation (eTable6 in Supplement2, eFigures2-5 in Supplement1).

Sensitivity analyses examining robustness of results to reasonable variation in participant inclusion criteria and outcome definitions were highly consistent with the primary analyses, with most proteins showing concordant effect estimates and directions. Using unimputed protein data instead of imputed values used in the primary analyses, we identified 1,576 proteins associated with at least one psychiatric condition (prevalent and/or incident) across all covariate adjustment levels, a net increase of 31 proteins (66 added, 35 lost) from primary analyses. Including participants with CRP levels >10 mg/L or missing values, previously excluded from the primary analysis, resulted in 1,693 proteins associated with at least one psychiatric condition, a net increase of 148 proteins (177 added, 29 lost). Excluding individuals who self-reported depression or anxiety at baseline identified an additional 184 proteins passing the significance thresholds, while 53 proteins no longer met these criteria. Across primary and sensitivity analyses, 1,685 proteins were associated with at least one psychiatric disorder (full results in eTable7 in Supplement2).

Plasma proteomics also captured disease-relevant signals in the negative and positive control conditions (eTable8 in Supplement2). Rheumatoid arthritis was associated with 779 proteins, dominated by immune markers (the strongest associations were observed for tumour necrosis factor (TNF), ribonucleoside-diphosphate reductase subunit M2 (RRM2), and C-C motif chemokine 7 (CCL7)). Senile cataract was associated with only 30 proteins, led by the lens protein β-crystallin B2 (CRYBB2), the sole protein associated with refraction and accommodation disorders.

Pathway enrichment analyses

Several biologically coherent pathways reached nominal significance (p<0.05) in Reactome and KEGG pathway enrichment analyses, although none survived FDR correction. Enrichment analyses for the shared proteins, associated with at least three disorders in fully adjusted models, identified immune-inflammatory signalling as a putative common mechanism for major psychiatric disorders (eFigure6 in Supplement1). Full enrichment results are presented in eTables9-10 in Supplement2.

Enrichment analyses for proteins associated with incident versus prevalent disorder were conducted for depression (eFigures7-8 in Suppplement1), as we had enough proteins in the fully adjusted models for depression but not the other disorders. These results indicate that pathway enrichment in incident depression is dominated by immune activation-related processes and inflammatory disease such as viral infection, inflammatory bowel disease, and rheumatoid arthritis, although the analyses did not account for other diagnoses preceding the depression diagnosis. In contrast, established depression was characterised by broader alterations in extracellular matrix, vascular, and metabolic signalling pathways. Enrichment analyses for proteins associated with other prevalent or incident disorders are presented in eFigures9-10 in Suppplement1.

Genetic causal inference using MR and colocalisation

To identify potentially causal proteins, we performed Mendelian randomisation analyses for 1,685 proteins identified across the primary and sensitivity analyses in UK Biobank, testing their genetically predicted effects on anxiety, depression, bipolar disorder, and schizophrenia using GWAS summary statistics for these disorders. For these 1,685 proteins, we extracted 1,691 GWASs (N=34,557, randomly selected sample of European-ancestry UKB participants with proteomics data), as GWAS data for CXCL8, IL6, LMOD1, SCRIB, and TNF were available across multiple Olink panels. In total, 3,830 blood protein quantitative trait loci (pQTLs) were selected as genetic instruments

In primary genetic analyses, we identified 125 protein-psychiatric disorder pairs (90 plasma proteins) with MR estimates surviving multiple-testing correction at FDR <0.05. The number of potentially causal proteins differed between disorders, with the highest number observed for psychosis (n=55), followed by bipolar disorder (n=22), depression (n=20), and anxiety (n=18). Of these, 32 pairs (26 proteins) showed genetic colocalisation (PP.H4 ≥0.8), passed Steiger filtering, and showed absence of heterogeneity and horizontal pleiotropy (where testable). 22 of these 26 proteins were instrumented by a single genetic variant.

For depression, five proteins fulfilled the predefined criteria for causality: cis-only instrumented SEMA3F (1 SNP; FDR=6.3×10^-4^), DDR1 (1 SNP; FDR=0.03), LTB (1 SNP; FDR=0.02), and trans-only BTN2A1 (1 SNP; FDR=4.9×10^-6^) and PAPPA (1 SNP; FDR=0.01). For bipolar disorder, six proteins met the criteria: *cis*-only CD40 (2 SNPs; FDR=0.05), CX3CL1 (1 SNP; FDR=0.01), DSG3 (1 SNP; FDR=0.01), PBXIP1 (1 SNP; FDR=0.02), and PI3 (1 SNP; FDR=0.01), and trans-only BTN2A1 (1 SNP; FDR=4.1×10^-5^).For psychotic disorder, 17 proteins satisfied criteria for causality (approximately 3 times the number of proteins linked to any other psychiatric diagnosis): cis-only instrumented AGER (1 SNP; FDR=1.7×10^-6^), C2 (1 SNP; FDR=5.4×10^-15^), NPTXR (1 SNP; FDR=0.002), ADAM22 (2 SNPs; FDR=0.01), CAPS (2 SNPs; FDR=0.005), CD40 (2 SNPs; FDR=0.04), CDHR1 (1 SNP; FDR=0.02), DDR1 (1 SNP; FDR=0.02), DNER (3 SNPs; FDR=0.006), PDIA3 (1 SNP; FDR=0.003) and RABEP1 (1 SNP; FDR=0.01), alongside trans-only BTN2A1 (1 SNP; FDR=4.7×10^-15^), MYBPC1 (1 SNP; FDR=0.02), MYL3 (1 SNP; FDR=0.02), MYLPF (1 SNP; FDR=0.03), MYOM3 (1 SNP; FDR=0.01) and SCG2 (1 SNP; FDR=0.006). For anxiety, four proteins satisfied the criteria. *cis*-only instrumented CD8A (SNP=1; FDR=0.002) and DDR1 (SNP=1; FDR=0.04), and trans-only BTNA2A1 (SNP=1; FDR=0.002) and VSIG4 (SNP=1; FDR=0.01).

In reverse-direction analyses, genetic liability to depression showed causal effects on six plasma proteins after Bonferroni correction (N tests=6,764; p<7.4×10^-6^), none overlapping with the 26 primary proteins. These 6 proteins were CXCL17 (P=1.7×10^-7^), ALPP (P=1.5×10^-6^), RARRES2 (P=1.9×10^-6^), PIGR (P=2.6×10^-6^), PRSS8 (P=6.6×10^-6^) and MSLN (P=7.1×10^-6^). We observed no convincing evidence that genetic liability to schizophrenia, bipolar disorder or anxiety causally influenced protein abundance after Bonferroni correction.

Findings remained largely consistent in sensitivity analyses using depression and bipolar disorder GWAS excluding UKB participants. Genetic instruments are detailed in eTables11-12 (Supplement2), and primary MR and colocalisation results in eTables13-15 (Supplement2).

In validation genetic analyses of the 26 primary proteins across the four psychiatric disorders, five protein-disorder pairs showed supportive causal evidence in two-sample MR using rare or ultrarare genetic variants, while 10 pairs received additional support from brain and blood gene expression (eQTL) MR and colocalisation analyses. In rare and ultrarare variant MR analyses, supportive causal estimates were observed for rare variant instrumented DDR1 on anxiety disorders (OR 1.19; 95%CI 1.05-1.35) and major depressive disorder (OR 1.42; 95% CI 0.98-2.08), CD40 on major depressive disorder (OR 0.54; 95%CI 0.39-0.75), and PBXIP1 on bipolar disorder (OR 0.46; 95%CI 0.23-0.91), as well as an ultrarare variant instrumented DSG3 on major depressive disorder (OR 0.38; 95%CI 0.17-0.83). In MR and genetic colocalisation using brain and blood eQTLs, there was evidence of genetically predicted gene expression of *CD40* in the brain cortex on schizophrenia and bipolar disorder, *DDR1* on anxiety*, DSG3* on bipolar disorder, and *SEMA3F* on depression. Additionally, there was evidence of blood genetically predicted gene expression of *ADAM22*, *CDHR1*, and *PDIA3* on schizophrenia, and of *AGER* on schizophrenia and bipolar disorder.

Primary and validation genetic results are summarised in Figure 2 (and in an expanded version with cross-sectional and longitudinal UKB associations in eFigure11 in Supplement1).  Validation instruments and results are detailed in eTables16-20 (Supplement2).

Appraisal

All 32 primary protein-disorder pairs (26 proteins) were appraised by instrument tier, validation score, and final evidence band (Table 2, eTable21 in Supplement2). Of these, 9 pairs (7 proteins) had supporting evidence for a plausible causal association and were classified as “Promising,” with their primary MR estimates and variant counts detailed in Table 3.

CD40 was the top-scoring protein for schizophrenia and bipolar disorder, instrumented by two independent non-MHC *cis* variants (Tier A). Higher genetically predicted CD40 levels were associated with lower risk of schizophrenia (OR 0.90; 95%CI 0.85-0.95) and bipolar disorder (OR 0.89; 95%CI 0.84-0.95), supported by concordant brain cortex eQTL MR with colocalisation for schizophrenia (OR 0.89; 95%CI 0.84-0.94) and bipolar disorder (OR 0.88; 95%CI 0.83-0.93), and concordant rare-variant MR for depression (OR 0.54; 95%CI 0.40-0.74).

Five additional pairs with Tier A instruments were classified as “Promising” from a single validation layer scoring +2: ADAM22, CDHR1, and PDIA3 for schizophrenia (colocalised blood eQTL MR), and SEMA3F for depression and DSG3 for bipolar disorder (colocalised brain cortex eQTL MR). Two pairs instrumented by a single MHC cis-variant (Tier B), DDR1 for anxiety and depression, were also classified as “Promising” via concordant rare-variant MR (less vulnerable than common variants to the attribution ambiguity in the MHC), with DDR1-anxiety also supported by colocalised brain cortex eQTL MR.

The remaining 23 pairs were classified as “Uncertain” due to non-significant or untestable validation layers, except for two directly contradicted pairs: AGER for schizophrenia (by colocalised blood eQTL MR) and PBXIP1 for bipolar disorder (by rare-variant MR).

Discovery, replication, and target characterisation

Four of the nine promising pairs (CD40, ADAM22, CDHR1, and PDIA3, each for schizophrenia) replicated a prior proteome-wide MR and colocalisation study with closely matching estimates[51]. CD40 for bipolar disorder has not been reported in a disorder-specific MR and colocalisation analysis but was implicated in cross-disorder studies[52, 53]. No prior MR and colocalisation reports were found for SEMA3F (depression), DSG3 (bipolar disorder), or DDR1 (anxiety or depression) (eTables27-28 in Supplement2).

Among the promising targets, human Mendelian phenotypes link *ADAM22* to developmental and epileptic encephalopathy and *CD40* to monogenic immune syndromes, supporting neurodevelopmental and immune processes in psychosis (eTable22 in Supplement2). Mouse knockout phenotypes link *DDR1* to hyperactivity, supporting a role in behavioural regulation, and *SEMA3F* to behavioural and neurological abnormalities, including altered locomotor activity, impaired exploration, and abnormal sensory responses (eTable23 in Supplement2).

Of the 26 primary proteins, 17 are potentially or strongly druggable, and five have approved or clinical-stage drugs: CD40, LTB, and CX3CL1 (Phase II autoimmune trials), AGER (Phase III Alzheimer’s disease trial), and MYL3 (approved for cardiovascular indications) (eTables24-26 in Supplement2).

eFigures

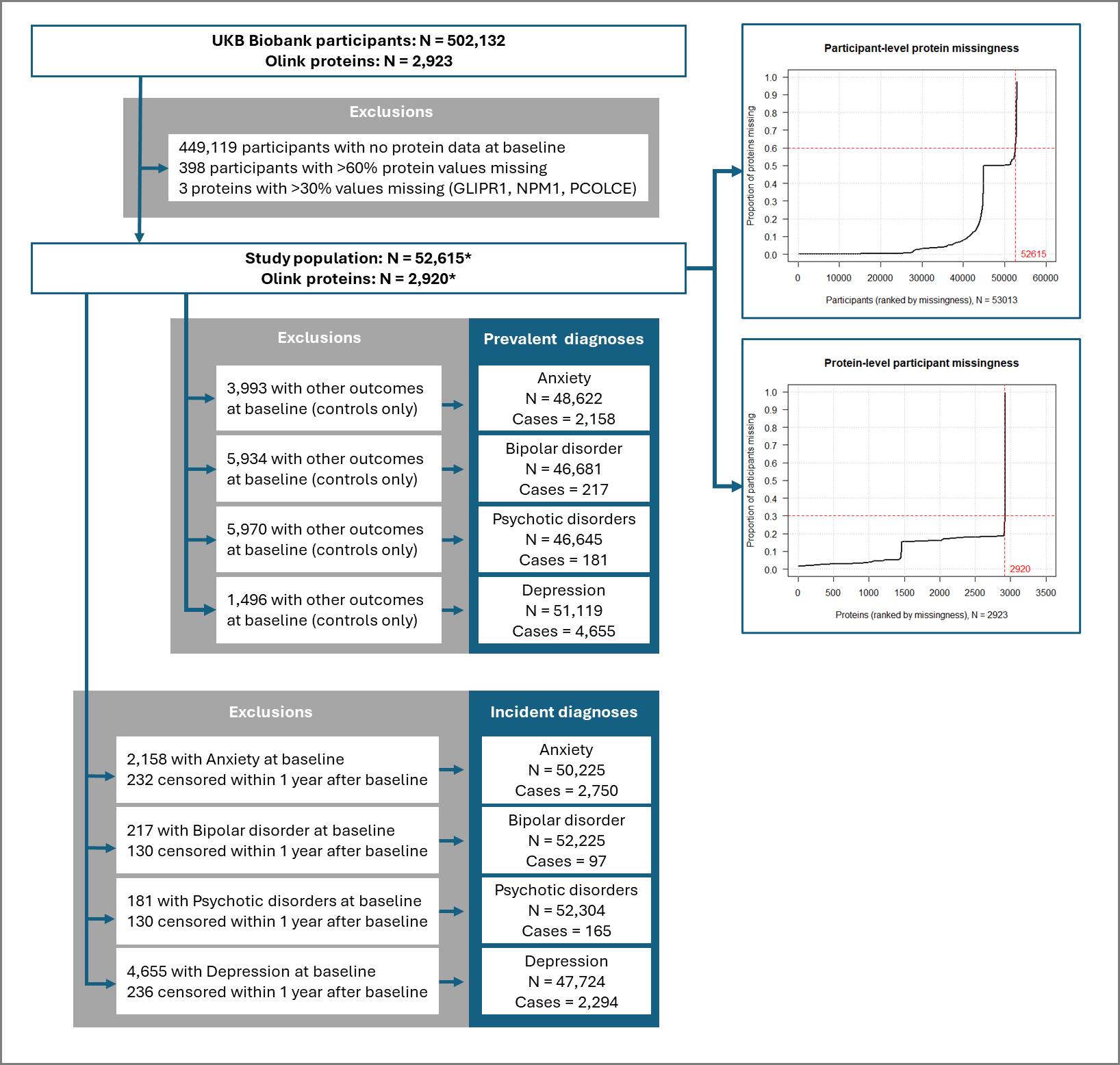

**eFigure1: Participant and protein inclusion and exclusion in UK Biobank cohort**

** Subplots show participant and protein missingness distributions with selection thresholds*

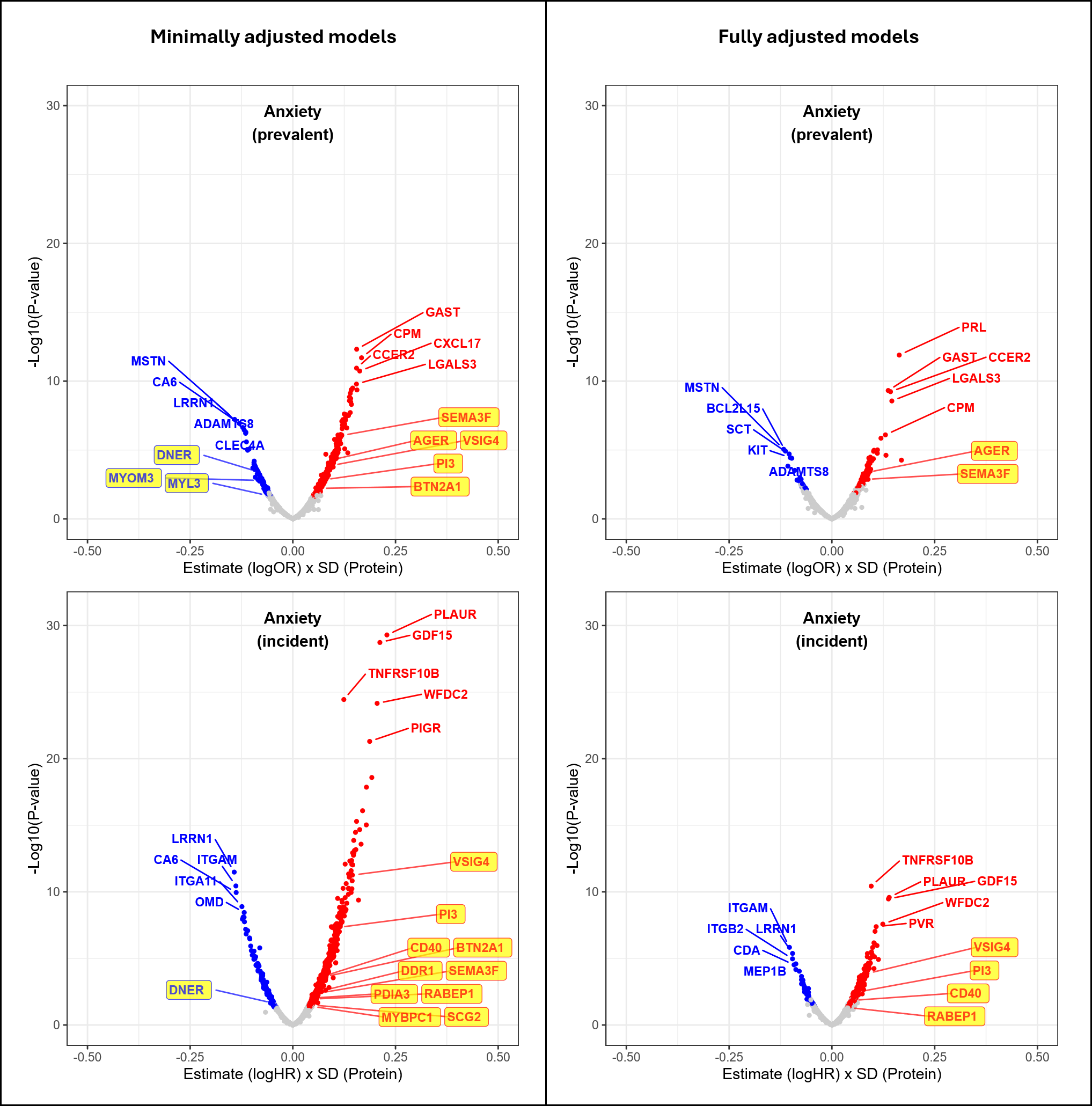

**eFigure2: Plasma proteomic associations with anxiety in UK Biobank.**

*Volcano plots showing the distribution, direction, and significance of protein-disorder associations from minimally and fully adjusted Cox (incident cases) and logistic (prevalent cases) regression models using imputed protein and covariate data. Participants with CRP levels at baseline above 10mg/L or missing were excluded. In incident analyses, participants censored within 12 months of baseline were excluded.*

*Protein-disorder associations passing the significance threshold (LFSR or FDR adjusted p-value < 0.05) are coloured according to direction of association, with positive associations shown in* ***red*** *and inverse associations in* ***blue****. All remaining associations are shown in* ***grey****. The five strongest positive and inverse associations (smallest P values) are annotated. Proteins with genetic evidence supporting causal effects for any of the four psychiatric disorders investigated (depression, anxiety, bipolar disorder, or psychotic disorders) are highlighted in* ***yellow****.*

*The y axis shows the −log10(p-value) value for each association. The x axis shows the standardised (per 1 SD increase in protein level) effect estimate, expressed as log HR (incident cases) or log OR (prevalent cases).*

*Covariate adjustment levels: Minimal = blood collection (season, time, fasting duration) + age + sex; Full = blood collection (season, time, fasting duration) + age + sex + Townsend Deprivation Index (TDI) + ethnicity + education + Body Mass Index (BMI) + physical activity + smoking + alcohol use.*

*CRP - C-Reactive Protein, FDR - False Discovery Rate, HR- Hazard Ratio, LFSR - Local False Sign Rate, OR - Odds Ratio.*

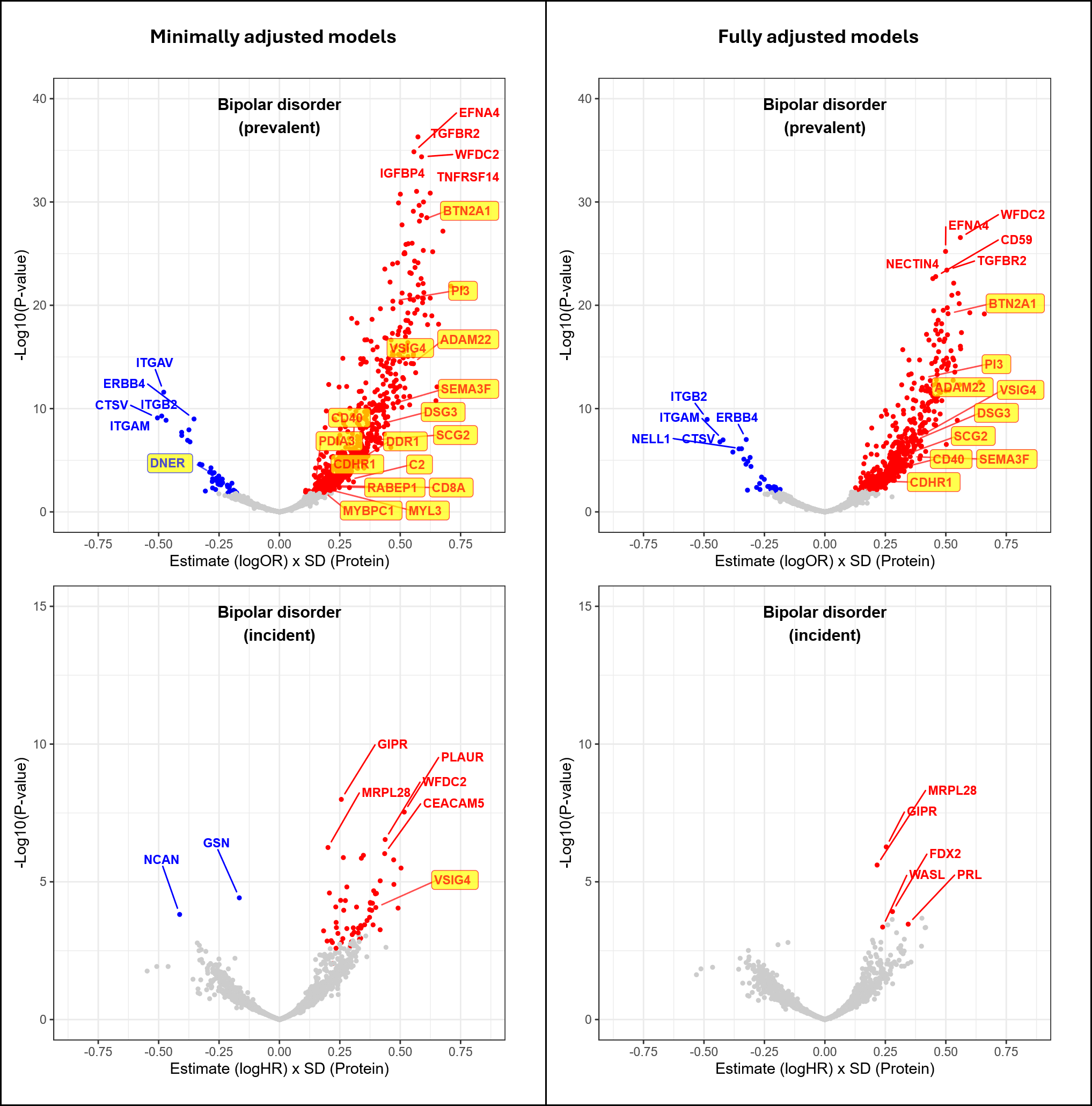

**eFigure3: Plasma proteomic associations with bipolar disorder in UK Biobank.**

*Volcano plots showing the distribution, direction, and significance of protein-disorder associations from minimally and fully adjusted Cox (incident cases) and logistic (prevalent cases) regression models using imputed protein and covariate data. Participants with CRP levels at baseline above 10mg/L or missing were excluded. In incident analyses, participants censored within 12 months of baseline were excluded.*

*Protein-disorder associations passing the significance threshold (LFSR or FDR adjusted p-value < 0.05) are coloured according to direction of association, with positive associations shown in* ***red*** *and inverse associations in* ***blue****. All remaining associations are shown in* ***grey****. The five strongest positive and inverse associations (smallest P values) are annotated. Proteins with genetic evidence supporting causal effects for any of the four psychiatric disorders investigated (depression, anxiety, bipolar disorder, or psychotic disorders) are highlighted in* ***yellow****.*

*The y axis shows the −log10(p-value) value for each association. The x axis shows the standardised (per 1 SD increase in protein level) effect estimate, expressed as log HR (incident cases) or log OR (prevalent cases).*

*Covariate adjustment levels: Minimal = blood collection (season, time, fasting duration) + age + sex; Full = blood collection (season, time, fasting duration) + age + sex + Townsend Deprivation Index (TDI) + ethnicity + education + Body Mass Index (BMI) + physical activity + smoking + alcohol use.*

*CRP - C-Reactive Protein, FDR - False Discovery Rate, HR- Hazard Ratio, LFSR - Local False Sign Rate, OR - Odds Ratio.*

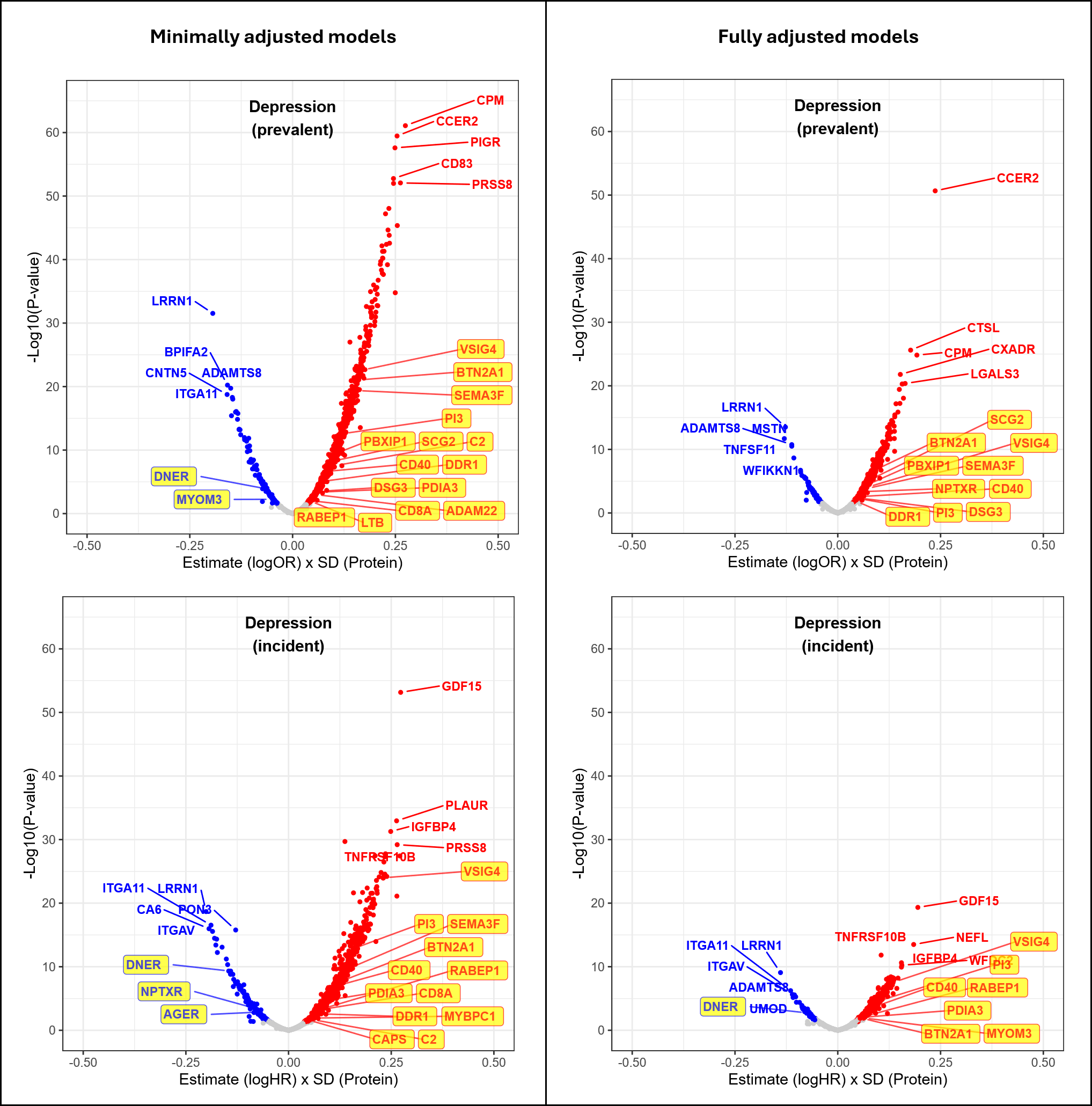

**eFigure4: Plasma proteomic associations with depression in UK Biobank.**

*Volcano plots showing the distribution, direction, and significance of protein-disorder associations from minimally and fully adjusted Cox (incident cases) and logistic (prevalent cases) regression models using imputed protein and covariate data. Participants with CRP levels at baseline above 10mg/L or missing were excluded. In incident analyses, participants censored within 12 months of baseline were excluded.*

*Protein-disorder associations passing the significance threshold (LFSR or FDR adjusted p-value < 0.05) are coloured according to direction of association, with positive associations shown in* ***red*** *and inverse associations in* ***blue****. All remaining associations are shown in* ***grey****. The five strongest positive and inverse associations (smallest P values) are annotated. Proteins with genetic evidence supporting causal effects for any of the four psychiatric disorders investigated (depression, anxiety, bipolar disorder, or psychotic disorders) are highlighted in* ***yellow****.*

*The y axis shows the −log10(p-value) value for each association. The x axis shows the standardised (per 1 SD increase in protein level) effect estimate, expressed as log HR (incident cases) or log OR (prevalent cases).*

*Covariate adjustment levels: Minimal = blood collection (season, time, fasting duration) + age + sex; Full = blood collection (season, time, fasting duration) + age + sex + Townsend Deprivation Index (TDI) + ethnicity + education + Body Mass Index (BMI) + physical activity + smoking + alcohol use.*

*CRP - C-Reactive Protein, FDR - False Discovery Rate, HR- Hazard Ratio, LFSR - Local False Sign Rate, OR - Odds Ratio.*

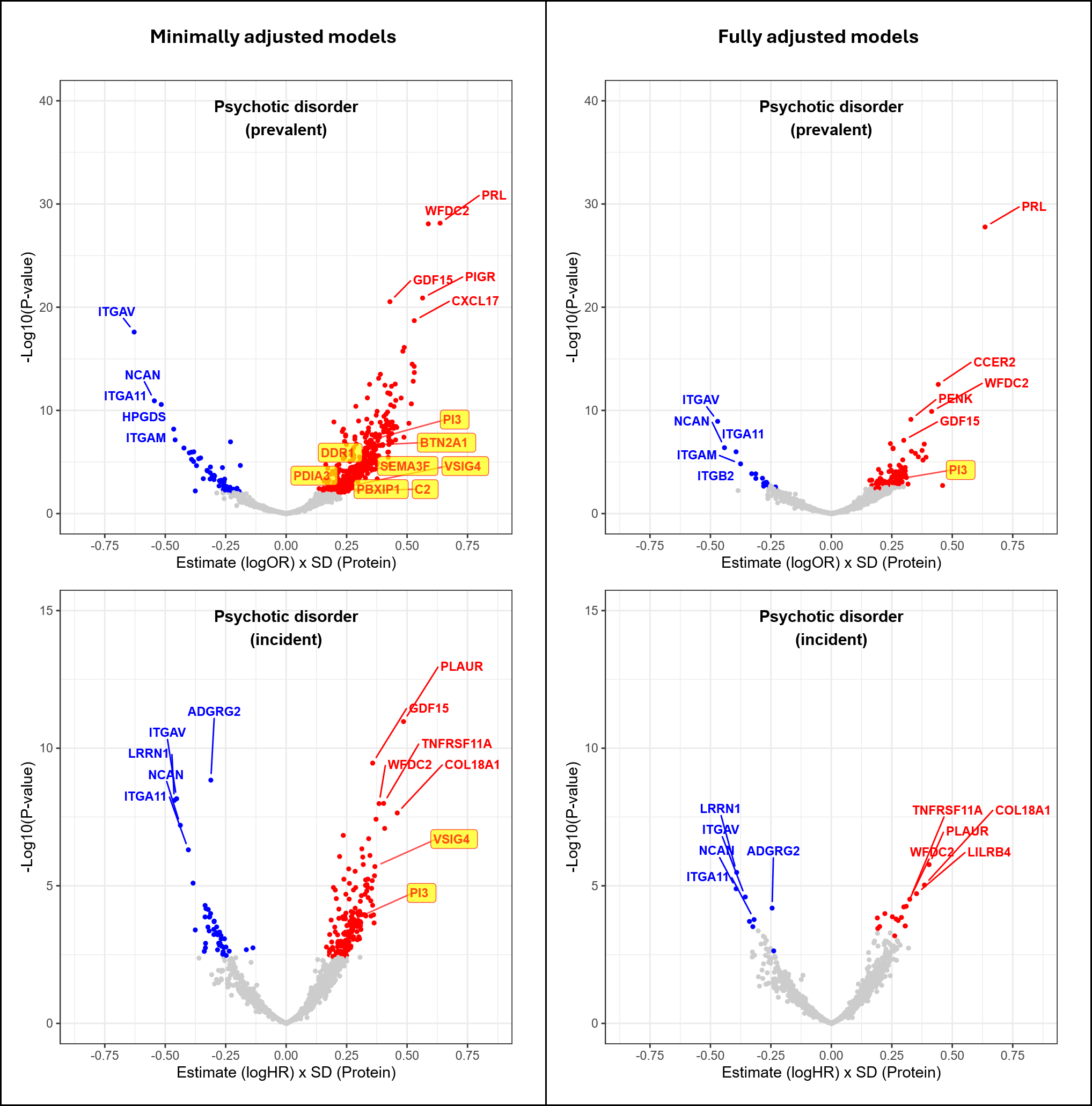

**eFigure5: Plasma proteomic associations with psychotic disorder in UK Biobank.**

*Volcano plots showing the distribution, direction, and significance of protein-disorder associations from minimally and fully adjusted Cox (incident cases) and logistic (prevalent cases) regression models using imputed protein and covariate data. Participants with CRP levels at baseline above 10mg/L or missing were excluded. In incident analyses, participants censored within 12 months of baseline were excluded.*

*Protein-disorder associations passing the significance threshold (LFSR or FDR adjusted p-value < 0.05) are coloured according to direction of association, with positive associations shown in* ***red*** *and inverse associations in* ***blue****. All remaining associations are shown in* ***grey****. The five strongest positive and inverse associations (smallest P values) are annotated. Proteins with genetic evidence supporting causal effects for any of the four psychiatric disorders investigated (depression, anxiety, bipolar disorder, or psychotic disorders) are highlighted in* ***yellow****.*

*The y axis shows the −log10(p-value) value for each association. The x axis shows the standardised (per 1 SD increase in protein level) effect estimate, expressed as log HR (incident cases) or log OR (prevalent cases).*

*Covariate adjustment levels: Minimal = blood collection (season, time, fasting duration) + age + sex; Full = blood collection (season, time, fasting duration) + age + sex + Townsend Deprivation Index (TDI) + ethnicity + education + Body Mass Index (BMI) + physical activity + smoking + alcohol use.*

*CRP - C-Reactive Protein, FDR - False Discovery Rate, HR- Hazard Ratio, LFSR - Local False Sign Rate, OR - Odds Ratio.*

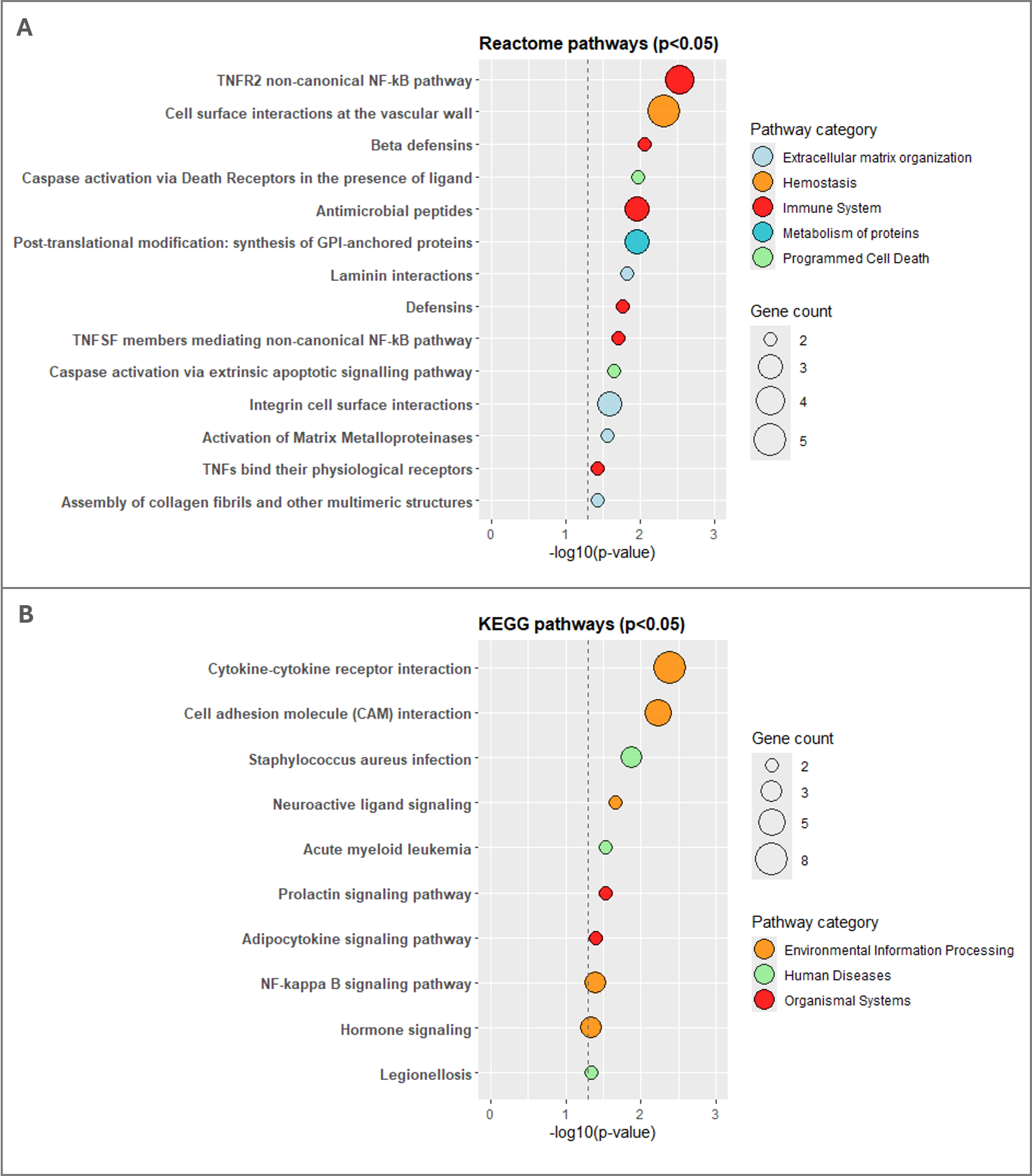

**eFigure6: Pathway enrichment for proteins associated with at least three out of four psychiatric disorders**

*A. Reactome pathways: Total = 118, Enriched (FDR < 0.05) = 0, Enriched (p-value < 0.05) = 14. B. KEGG pathways: Total = 75, Enriched (FDR < 0.05) = 0, Enriched (p-value < 0.05) = 10. Psychiatric disorders: anxiety, bipolar disorder, depression, psychotic disorder.*

*Genes encoding proteins (Proteins = 37) passing significance threshold (LFSR or FDR < 0.001) in Cox (incident) or logistic (prevalent) regression models were included in enrichment analyses. Genes encoding 2,920 UKB proteins served as the background gene set.*

*Covariate adjustment level: Full = blood collection (season, time, fasting duration) + age + sex + Townsend Deprivation Index (TDI) + ethnicity + education + Body Mass Index (BMI) + physical activity + smoking + alcohol use.*

*The y axis indicates the −log10(p-value) values for enrichment, with threshold of -log10(0.05) displayed. The x axis displays enrichment terms stratified by top level pathway category. Only pathways with p-value >0.05 are shown.*

*FDR - False Discovery Rate, KEGG - Kyoto Encyclopaedia of Genes and Genomes, LFSR - Local False Sign Rate, UKB - United Kingdom Biobank.*

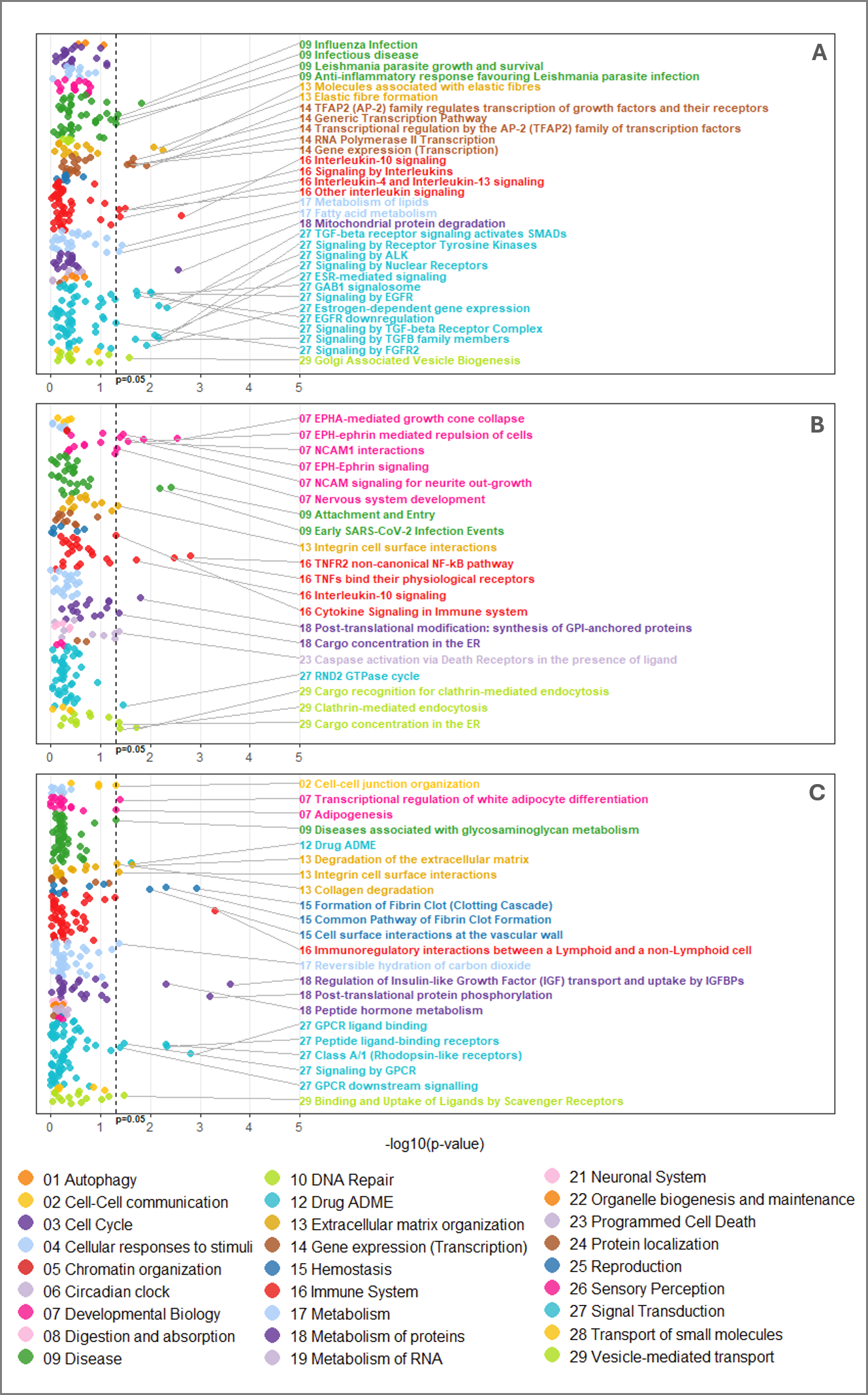

**eFigure7: Reactome pathway enrichment for proteins associated with depression (UK Biobank)**

*A. Proteins associated with incident depression only (not prevalent depression): Proteins = 120; Pathways: Total = 324, Enriched (FDR <0.05) = 0, Enriched (p-value <0.05) = 31.*

*B. Proteins associated with both incident and prevalent depression: Proteins = 83; Pathways: Total = 219, Enriched (FDR <0.05) = 0, Enriched (p-value <0.05) = 19.*

*C. Proteins associated with prevalent depression only (not incident depression): Proteins = 221; Pathways: Total = 341, Enriched (FDR <0.05) = 0, Enriched (p-value <0.05) = 22.*

*Incident depression refers to diagnoses recorded 12 months after the UK Biobank baseline assessment, and prevalent depression refers to lifetime diagnoses recorded prior to baseline.*

*Genes encoding proteins passing significance threshold (LFSR or FDR < 0.001) in Cox (incident) or logistic (prevalent) regression models were included in enrichment analyses. Genes encoding 2,920 UKB proteins served as the background gene set.*

*Covariate adjustment level: Full = blood collection (season, time, fasting duration) + age + sex + Townsend Deprivation Index (TDI) + ethnicity + education + Body Mass Index (BMI) + physical activity + smoking + alcohol use.*

*The y axis indicates the −log10(p-value) values for enrichment, with threshold of -log10(0.05) displayed. The x axis displays enrichment terms stratified by top level pathway category.*

*FDR - False Discovery Rate, KEGG - Kyoto Encyclopaedia of Genes and Genomes, LFSR - Local False Sign Rate, UKB - United Kingdom Biobank.*

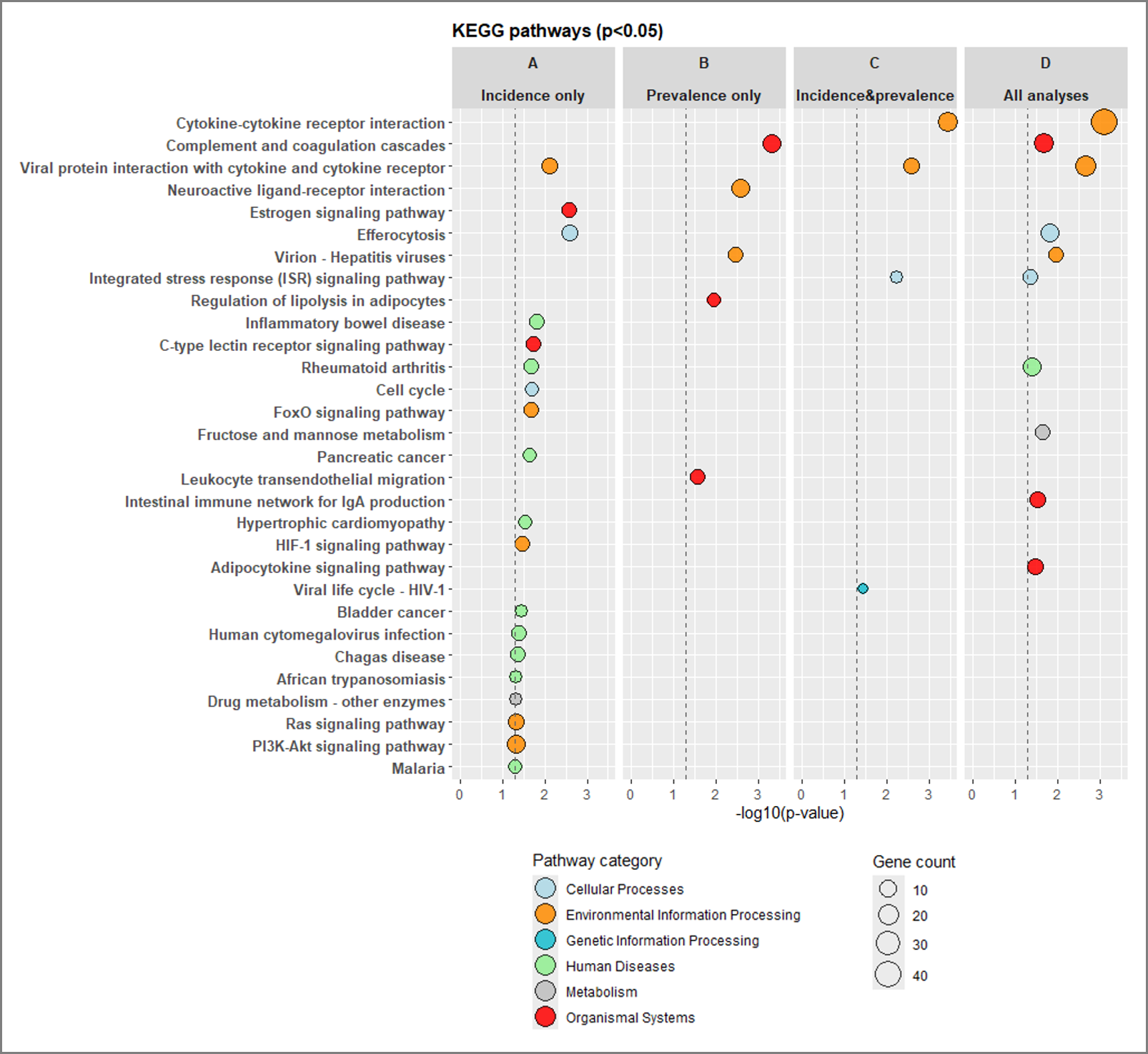

**eFigure8: KEGG pathway enrichment for proteins associated with depression (UK Biobank)**

*A. Incidence only: Proteins associated with incident depression only (not prevalent depression): Proteins = 120; Pathways: Total = 190, Enriched (FDR <0.05) = 0, Enriched (p-value <0.05) = 10.*

*B. Incidence and prevalence: Proteins associated with both incident and prevalent depression: Proteins = 83; Pathways: Total = 111, Enriched (FDR <0.05) = 1, Enriched (p-value <0.05) = 4.*

*C. Prevalence only: Proteins associated with prevalent depression only (not incident depression): Proteins = 221; Pathways: Total = 179, Enriched (FDR <0.05) = 0, Enriched (p-value <0.05) = 5.*

*D. All analyses: Proteins associated with incident or prevalent depression: Proteins=424; Pathways: Total = 232, Enriched (FDR <0.05) = 0, Enriched (p-value <0.05) = 10.*

*Incident depression refers to diagnoses recorded 12 months after the UK Biobank baseline assessment, and prevalent depression refers to lifetime diagnoses recorded prior to baseline. The y axis indicates the −log10(p-value) values for enrichment, with threshold of -log10(0.05) displayed. The x axis displays enrichment terms stratified by top level pathway category. Only pathways with p-value >0.05 are shown.*

*Genes encoding proteins passing significance threshold (LFSR or FDR <0.001) in Cox (incident) or logistic (prevalent) regression models were included in enrichment analyses. Genes encoding 2,920 UKB proteins served as the background gene set. Covariate adjustment level: Full = blood collection (season, time, fasting duration) + age + sex + Townsend Deprivation Index (TDI) + ethnicity + education + Body Mass Index (BMI) + physical activity + smoking + alcohol use.*

*FDR - False Discovery Rate, KEGG- Kyoto Encyclopaedia of Genes and Genomes, LFSR - Local False Sign Rate, UKB - United Kingdom Biobank*

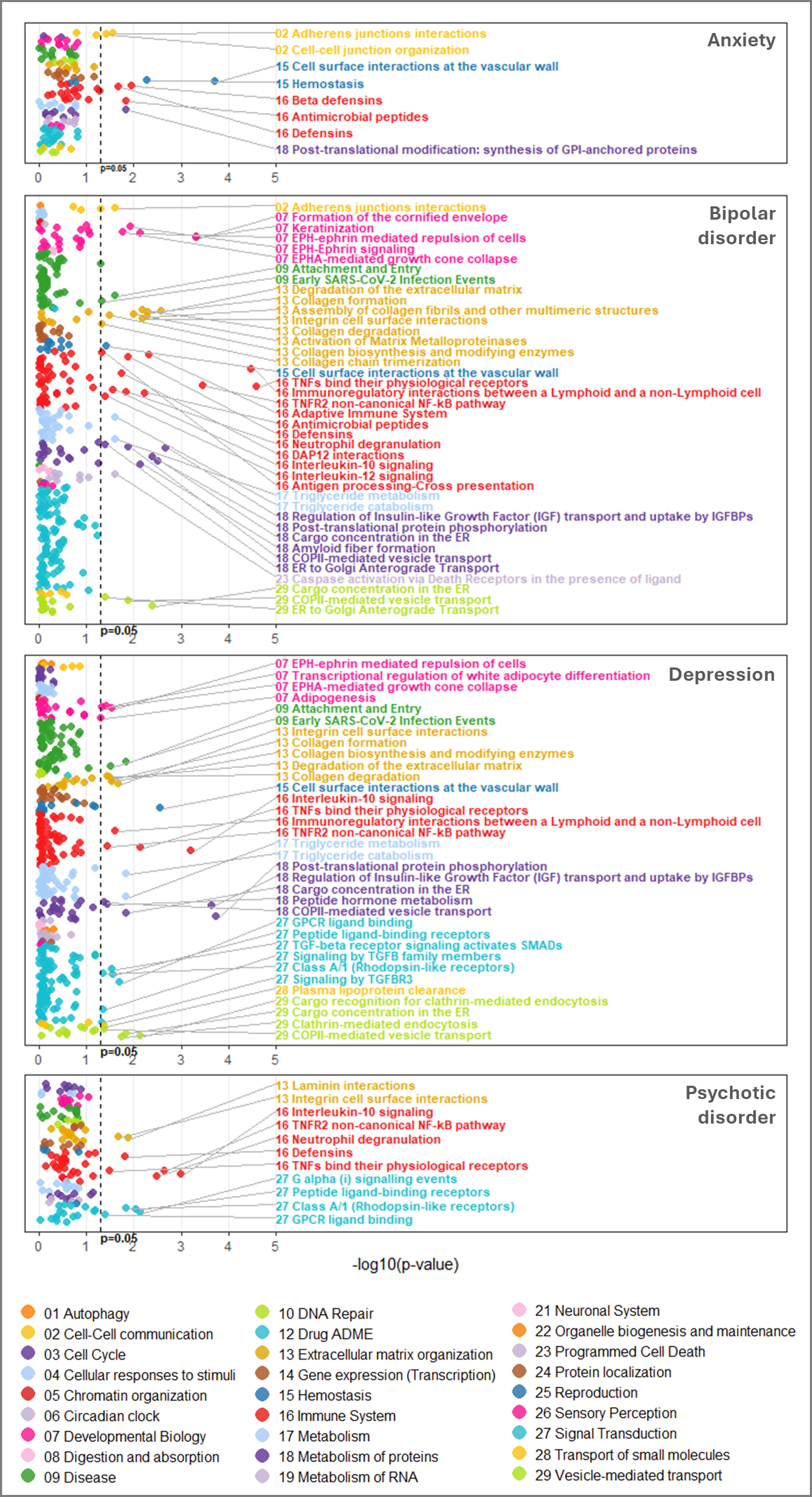

**eFigure9: Reactome pathway enrichment for proteins associated with psychiatric disorders (fully adjusted models)**

*Proteins associated with incident or prevalent anxiety: Proteins = 45; Pathways: Total = 126, Enriched (FDR <0.05) = 1, Enriched (p-value <0.05) = 8.*

*Proteins associated with incident or prevalent bipolar disorder: Proteins = 323; Pathways: Total = 382, Enriched (FDR <0.05) = 5, Enriched (p-value <0.05) = 37.*

*Proteins associated with incident or prevalent depression: Proteins = 424; Pathways: Total = 477, Enriched (FDR <0.05) = 0, Enriched (p-value <0.05) = 31.*

*Proteins associated with incident or prevalent psychotic disorder: Proteins = 38; Pathways: Total = 149, Enriched (FDR <0.05) = 0, Enriched (p-value <0.05) = 11.*

*Genes encoding proteins passing significance threshold (LFSR or FDR < 0.001) in Cox (incident) or logistic (prevalent) regression models were included in enrichment analyses. Genes encoding 2,920 UKB proteins served as the background gene set.*

*Covariate adjustment level: Full = blood collection (season, time, fasting duration) + age + sex + Townsend Deprivation Index (TDI) + ethnicity + education + Body Mass Index (BMI) + physical activity + smoking + alcohol use.*

*The y axis indicates the −log10(p-value) values for enrichment, with threshold of -log10(0.05) displayed. The x axis groups enrichment terms by top level pathway category.*

*FDR - False Discovery Rate, LFSR - Local False Sign Rate, UKB - United Kingdom Biobank.*

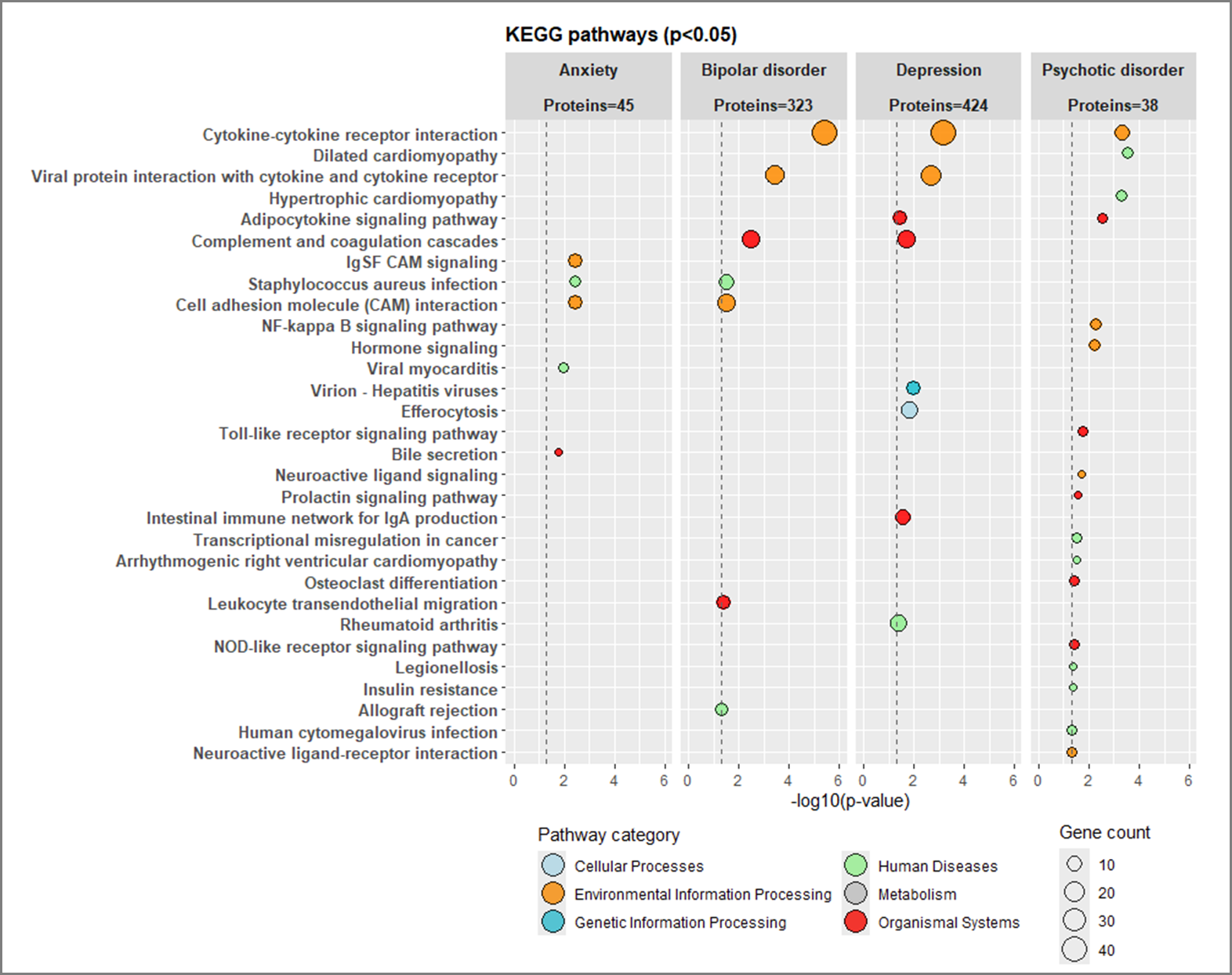

**eFigure10: KEGG pathway enrichment for proteins associated with psychiatric disorders (fully adjusted models)**

*Proteins associated with incident or prevalent anxiety: Proteins = 45; Pathways: Total = 86, Enriched (FDR <0.05) = 1, Enriched (p-value <0.05) = 5.*

*Proteins associated with incident or prevalent bipolar disorder: Proteins = 323; Pathways: Total = 185, Enriched (FDR <0.05) = 2, Enriched (p-value <0.05) = 7.*

*Proteins associated with incident or prevalent depression: Proteins = 424; Pathways: Total = 231, Enriched (FDR <0.05) = 0, Enriched (p-value <0.05) = 8.*

*Proteins associated with incident or prevalent psychotic disorder: Proteins = 38; Pathways: Total = 103, Enriched (FDR <0.05) = 3, Enriched (p-value <0.05) = 17.*

*Genes encoding proteins passing significance threshold (LFSR or FDR <0.001) in Cox (incident) or logistic (prevalent) regression models were included in enrichment analyses. Genes encoding 2,920 UKB proteins served as the background gene set.*

*Covariate adjustment level: Full = blood collection (season, time, fasting) + age + sex + Townsend Deprivation Index (TDI) + ethnicity + education + Body Mass Index (BMI) + physical activity + smoking + alcohol use.*

*The y axis indicates the −log10(p-value) values for enrichment, with threshold of -log10(0.05) displayed. The x axis groups enrichment terms by top level pathway category. Only pathways with p-value >0.05 are included.*

*FDR - False Discovery Rate, KEGG- Kyoto Encyclopaedia of Genes and Genomes, LFSR - Local False Sign Rate, UKB - United Kingdom Biobank.*

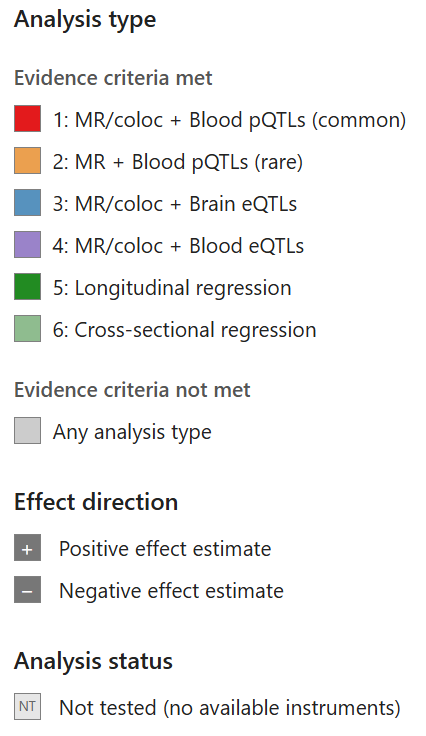

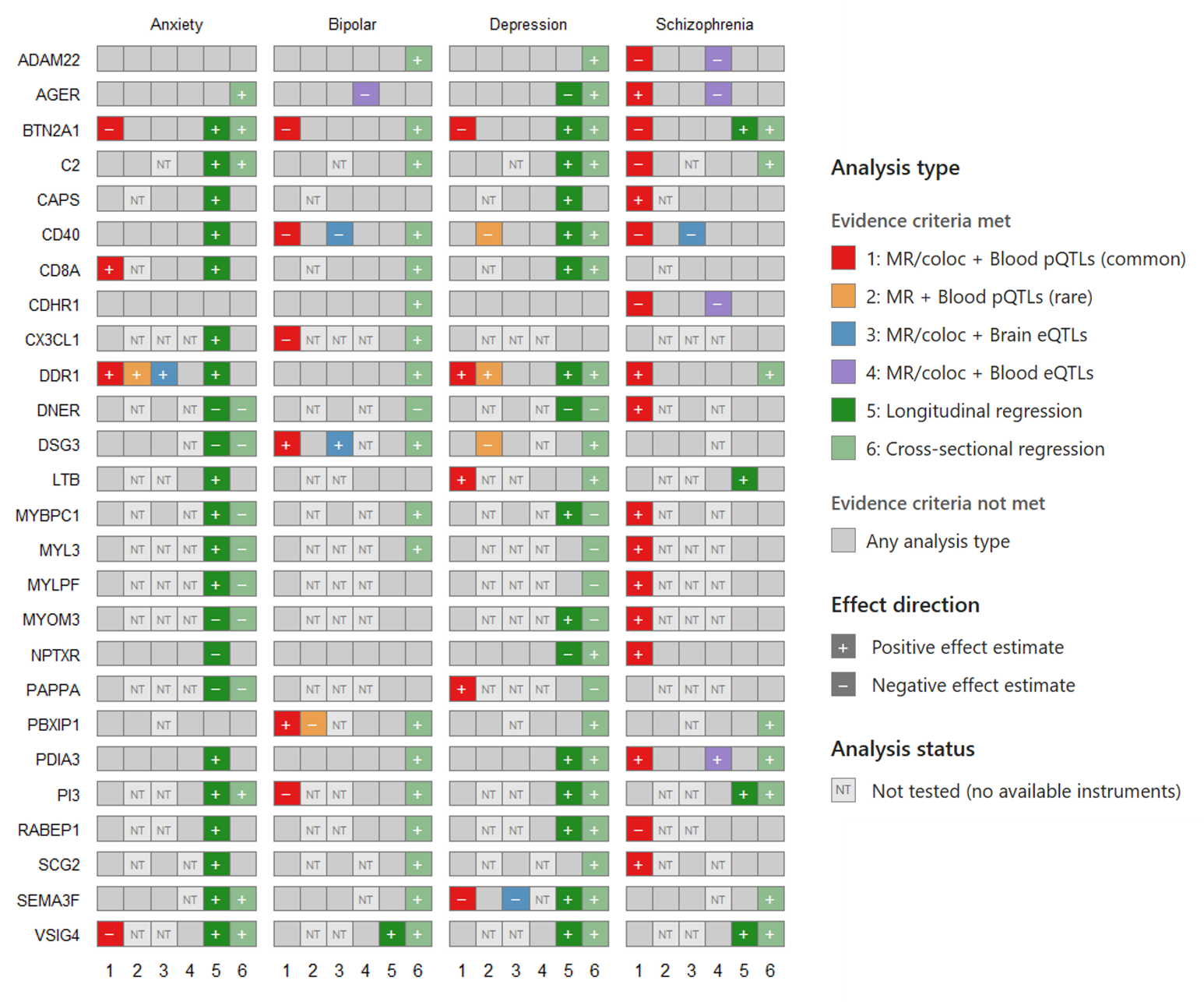

**eFigure11: Evidence for protein-disorder markers from primary and validation genetic analyses alongside evidence from cross-sectional and longitudinal analyses in UK Biobank**

*The primary analysis (1), using MR and genetic colocalisation with common-variant blood pQTLs (cis-only or trans-only), was considered to provide evidence for a protein-disorder association if it met all of the following criteria: MR FDR <0.05, genetic colocalisation PP.H4 ≥0.8, no evidence of heterogeneity or horizontal pleiotropy (where testable), passing Steiger filtering, and no evidence of reverse causation. 32 protein-disorder markers across 26 proteins met these criteria.*

*The validation analyses comprised (2) MR using rare or ultrarare variant blood pQTLs (cis-only), (3) MR and genetic colocalisation using brain cortex eQTLs (cis-only), and (4) MR and genetic colocalisation using blood eQTLs (cis-only). A validation analysis was considered to provide supporting evidence if the MR P value was <0.05 and, for eQTL-based analyses, genetic colocalisation PP.H4 was ≥0.8.*

*Results from the minimally and fully adjusted regression models in longitudinal (5) and cross-sectional (6) association analyses in UK Biobank (associations with incident or prevalent cases passing significance thresholds of FDR or LFSR < 0.05) are also included. Models with both imputed and unimputed protein data were included.*

*Effect direction was defined by the sign of the MR effect estimate or regression estimate (log odds ratio).*

*Abbreviations: coloc, colocalisation; eQTL, expression quantitative trait locus, FDR, false discovery rate; LFSR, local false sign rate; MR, Mendelian randomisation; NT, not testable; PP.H4, posterior probability of a shared causal variant; pQTL, protein quantitative trait locus.*
